# Causal relationship between liver fat reduction and clinical outcomes in human genetic studies and randomized clinical trials of metabolic-dysfunction associated steatohepatitis

**DOI:** 10.64898/2026.09.27.26364101

**Authors:** Niek Verweij, George Hindy, Quang Nguyen, Olukayode Sosina, Kusha A Mohammadi, Tejasv Bedi, Joseph Herman, Karl Landheer, Benjamin Geraghty, Selin Somersan-Karakaya, Adam Locke, Liron Ganel, Arthur Gilly, Rouel Lanche, Stefanie Hectors, Mary Germino, Gavin H. Imperato, Ifeanyi Anidi, Lauryn Choleva, Emily Labriola-Tompkins, Muhammad Effendi, Katie Tuckwell, Sonya Aziz-Zaman, Jane Witkin, Jutta L Miller, Tae-Hwi Schwantes-An, Regeneron Genetics Center, GHS-RGC DiscovEHR Collaboration, Penn Medicine Biobank, Colorado Center for Personalized Medicine, Mayo Clinic-RGC Project Generation, Malmö Diet and Cancer Study, Indiana University School of Medicine (Indiana-CLDB), UCLA-RGC ATLAS Collaboration, Mount Sinai Million Health Discoveries Program, Rohit Loomba, Naga P. Chalasani, Rong Liu, Mark Sleeman, Viktoria Gusarova, Gary A Herman, Jonathan Marchini, Aris Baras, Thomas D Norton, Luca A. Lotta, David J Lederer

## Abstract

Reliance on invasive liver biopsy endpoints is a major limiting factor in new therapeutic development for metabolic dysfunction-associated steatohepatitis (MASH). While liver fat accumulation is considered a driver of early disease, liver fat reduction is not recognized as a surrogate endpoint by health authorities. Here, we conducted the largest-to-date genome-wide association study (GWAS) of liver fat in 79,642 people with magnetic resonance imaging and developed a polygenic score with liver fat-lowering alleles across 55 genomic loci. In a separate set of 714,886 individuals, a 30% relative liver fat reduction due to the polygenic score was associated with 39% lower odds of liver cirrhosis and 41% lower odds of hepatocellular carcinoma, consistent with a strong causal relationship between lifelong liver fat differences and advanced liver clinical outcomes. A systematic review and meta-analysis of 20 randomized controlled trials including a total of 3,502 MASH patients showed that treatment-induced liver fat reductions were associated with MASH resolution (meta-regression slope in % units of the outcome for each 1% relative reduction in liver fat from baseline, β = -0.70%, p<0.001) and fibrosis improvement (β = -0.24%, p<0.001). These findings demonstrate the profound etiologic impact of liver fat in MASH and provide compelling evidence for liver fat reductions as a reasonably likely surrogate endpoint for future clinical development.

## Introduction

Metabolic dysfunction-associated steatohepatitis (MASH) is characterized by triglyceride accumulation in hepatocytes leading to lobular inflammation, hepatocyte injury, and consequent fibrosis, which can progress to cirrhosis and its complications.^1^ Approximately 30% (1.6 billion) of adults worldwide are estimated to have metabolic dysfunction associated liver disease (MASLD), a precursor of MASH that is defined by excess liver fat accumulation, and 5% (approximately 260 million) are estimated to have MASH.^2-4^ Despite intense effort, only two therapeutics for MASH have received conditional regulatory approval in the United States, leaving substantial unmet need in this patient population^5^.

The current paradigm for therapeutic development in MASH relies on demonstration of histological improvement to seek conditional regulatory approval and confirmation of improvement in clinical outcomes for full regulatory approval.^6,7^ This approach has led to prolonged drug development timelines and requires liver biopsies, which are associated with procedural complications including death and yield highly variable assessments by pathologists.^8^ The use of non-invasive reasonably likely surrogate endpoints could speed drug development and avoid problematic liver biopsies^9^, simultaneously reducing patient burden and increasing willingness to participate in clinical trials.

Change in liver fat is a candidate surrogate endpoint with strong face validity^10^. It is widely accepted that liver fat is the early initiator of the etiologic processes leading to MASH. However, there is limited experimental evidence that changes in liver fat impact clinical outcomes in MASH. We set out to generate data to (1) estimate the etiologic relationship between genetically-determined differences in liver fat and the risk of liver outcomes in the general population, (2) assess the prognostic role of liver fat for liver outcomes in the general population, and (3) estimate whether the magnitude of therapeutic-induced reductions in liver fat is associated with changes in histology endpoints in patients with MASH in randomized controlled trials.

## Results

### Genome-wide association study identifies novel loci for liver fat

We performed the largest-to-date genome-wide association study (GWAS) of liver fat in 79,642 UK Biobank (UKB) participants with available magnetic resonance imaging (MRI)-derived proton density fat fraction (PDFF), the accepted non-invasive gold standard for hepatic fat quantification^11^ (**Supplementary Table 1)**. We used REGENIE ^12^(ref) to fit linear whole-genome regression models to estimate the association of each of the 10,764,995 ascertained common variants (minor allele frequency>0.5%) with PDFF (**Methods**). Quality control statistics were consistent with optimal control of population stratification (linkage-disequilibrium score regression intercept value, 0.975; attenuation ratio, −0.02; estimated common-variants based heritability, 23%; **Supplementary Figure 1**), as expected for the REGENIE approach^12^. Common genetic variants at 56 loci reached the genome-wide level of statistical significance for association with PDFF (P<5×10^-8^, **Figure 1**, **Supplementary Table 2**), including 31 (55%) novel loci, not identified in previously published GWAS of liver fat measurements^13-19^.

**Figure 1.**
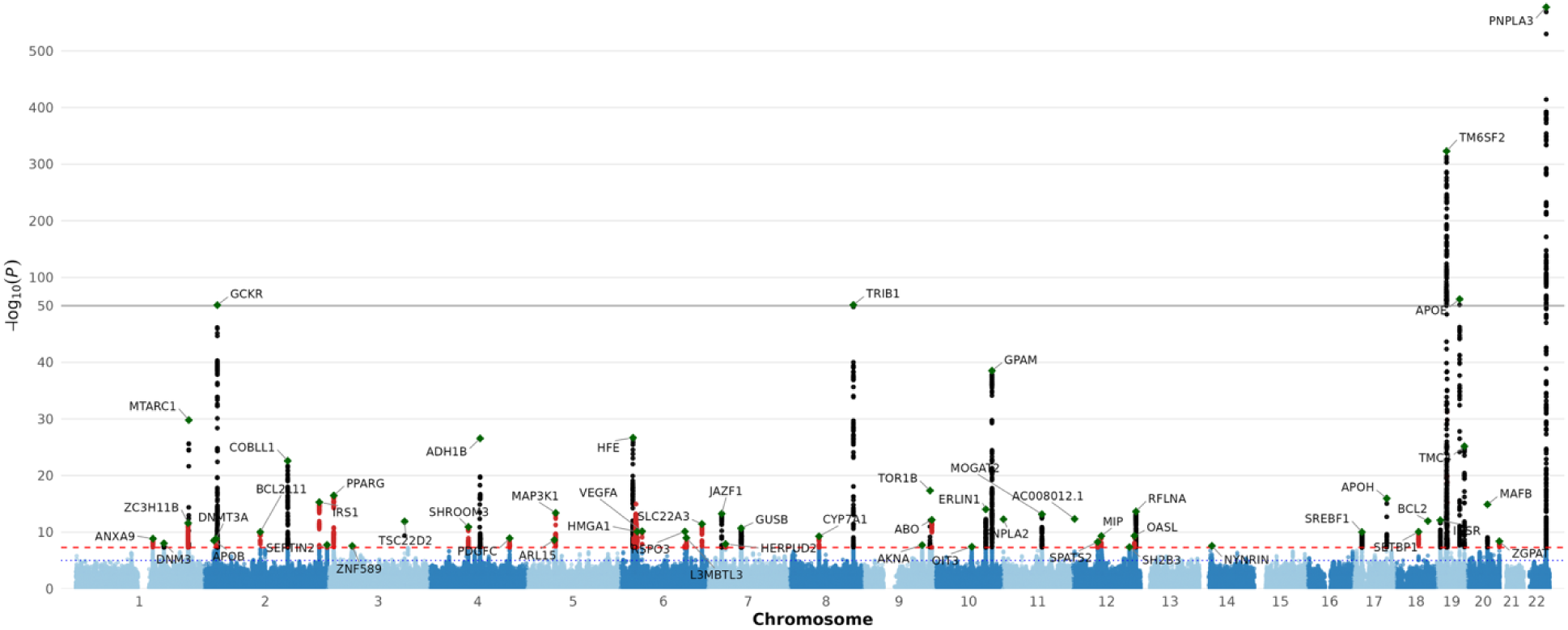
Manhattan plot for liver fat percentage measured by PDFF-MRI. In the largest genome-wide association study of liver fat to date, common genetic variants at 56 loci reached genome-wide significance (P < 5×10, red dotted line); for details see **Supplementary Table 2**. Green diamonds indicate the lead variant at each associated locus. Red dots represent genetic variants at novel loci not previously reported in association with liver fat; black dots represent variants at loci previously reported within 1 megabase of the loci identified in the current analysis. Genetic variants (dots) are arranged by genomic position on the x-axis, with the - log10(P-value) on the y-axis.

### Genetic evidence: Lower levels of liver fat due to genotype are strongly associated with protection against severe liver disease outcomes

To estimate the association with severe liver outcomes of lifelong differences in liver fat due to genotype, we generated a polygenic score (PRS) for lower liver fat using 55 of the 56 genomic loci implicated by our GWAS (excluding the *HFE* locus, which is primarily related to liver iron accumulation; **Methods**). The PRS was strongly associated with liver fat in an independent validation set of 10,377 UKB participants with PDFF who were not included in the GWAS (P = 6.1×10^-119^); each standard deviation (SD) increase in the PRS was associated with a 4.9-percentage-point increase in measured PDFF (95% CI: 4.5, 5.3%), equivalent to 0.93 SDs of measured PDFF (95% CI: 0.85, 1.01 SDs). We next estimated associations with liver cirrhosis in 15,112 cases and 699,774 controls from 10 cohorts, not included in our GWAS analysis. In this analysis, a 30% relative decrease (corresponding to -1.75 PDFF percentage units) in liver fat due to the 55 loci was associated with 39% lower odds of cirrhosis (OR = 0.61, 95% CI: 0.58 to 0.64; P=1x10^-81^; **Figure 2A**), with evidence of a strong log-linear relationship across individual variants). This association was robust to sensitivity analyses using alternative MR methods (MR-Egger, weighted median, penalised weighted median, simple median, and weighted mode), all of which yielded consistent effect estimates and directions of association (ORs ranging from 0.56 to 0.69 per 30% relative decrease in liver fat; **Supplementary Table 3**), as well as to leave-one-out analyses excluding one locus at a time (**Supplementary Figure 2)**. Of the 55 loci, 17 (30%) were individually associated with risk of cirrhosis (p<9×10^-4^, a Bonferroni correction for 55 loci - **Supplementary Table 4**) and 52 (95%) showed a directional consistency between association estimates for liver fat and cirrhosis (p_binomial_=7.7×10^-13^). In an individual-level analysis excluding related participants, we estimated the association with cirrhosis stratified by PRS quintiles. Cirrhosis risk rose monotonically across quintiles with evidence of a strong dose-response relationship (**Figure 2**).

**Figure 2:**
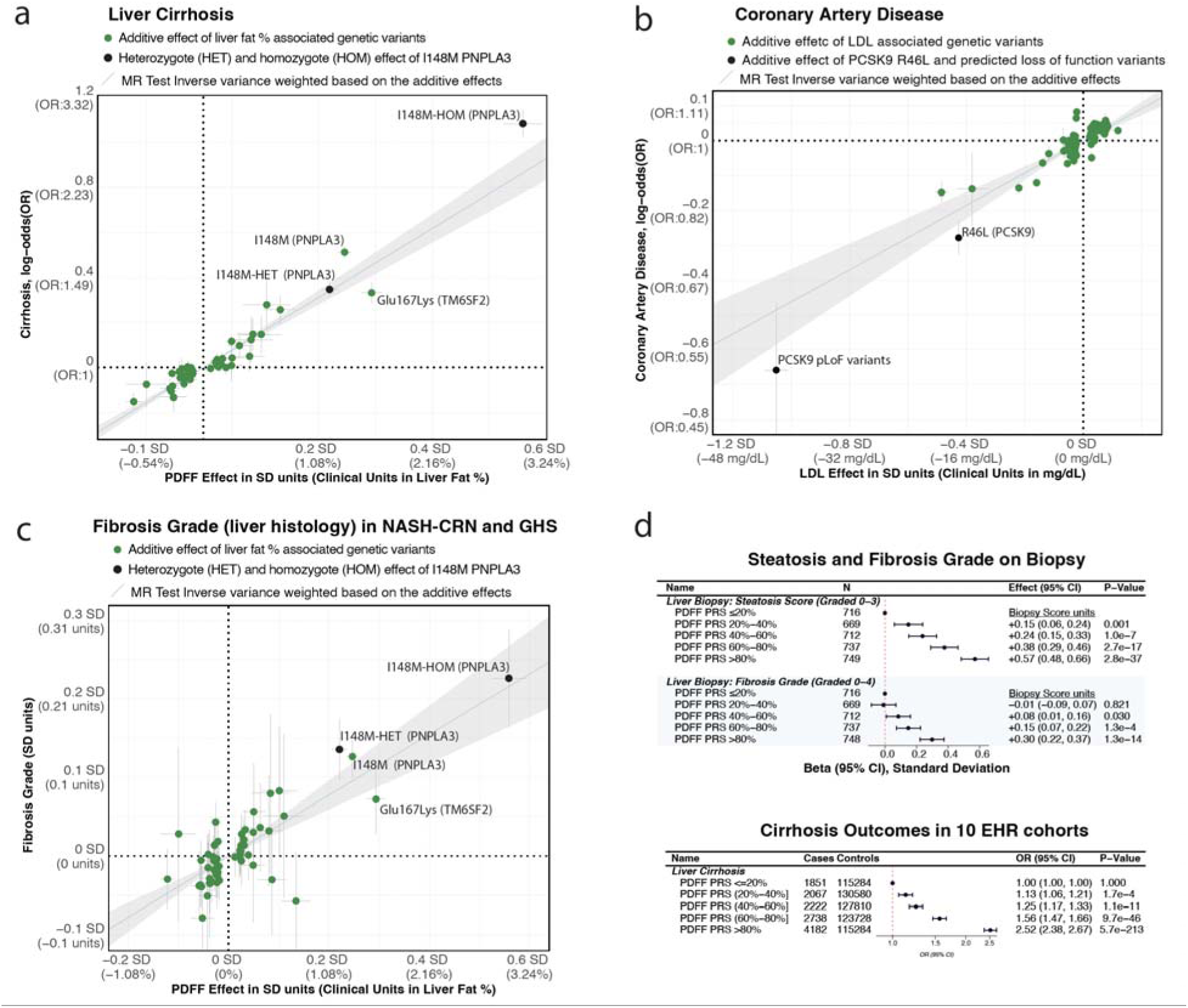
Mendelian randomization analyses: The Genetically Predicted Effect of liver fat Percentage by PDFF on Liver Cirrhosis is Strong and Comparable to the Genetically Predicted Effect of LDL-C on Coronary Artery Disease. A 30% decrease in genotype-predicted liver fat by PDFF is associated with 39% lower odds of cirrhosis (OR=0.61), a magnitude comparable to LDL-C and CAD, where a 30% LDL-C decrease also yields 39% lower odds of CAD (OR=0.61). Panels: (A) PDFF-cirrhosis (see also **Supplementary Table 3**); (B) LDL-C–CAD; (C) PDFF–fibrosis grade the GHS Bariatric and NASH-CRN cohorts; (D) association between individual level PDFF polygenic risk score and steatosis/fibrosis grades on biopsy in the GHS Bariatric cohort and Cirrhosis outcomes in 10 EHR cohorts.

The relationship between differences in liver fat due to genotype and the risk for hepatocellular carcinoma was as strong as that for cirrhosis; a 30% relative decrease in liver fat was associated with a 41% reduction in the odds of hepatocellular carcinoma (OR, 0.59; 95% CI, 0.55 to 0.62, p = 1x10^-71^). There were also strong associations with liver fibrosis score at liver histology in 7,250 participants with liver biopsy (**Figure 2, Supplementary Figure 3, Supplementary Table 3-4**).

We compared the association between liver fat and liver outcomes to the well-known causal relationship between low-density lipoprotein cholesterol (LDL-C) and coronary artery disease (CAD), where LDL-C reductions are accepted as a validated surrogate endpoint for full approval of lipid lowering agents. In 88,234 cases and 626,124 controls, a 30% decrease in LDL-C (corresponding to -43 mg/dL) was associated with a 39% lower odds of CAD (OR, 0.61; 95% CI, 0.55 to 0.68, P=6×10^-22^; **Figure 2B**), a near-identical relationship as that between liver fat and cirrhosis risk.

### Epidemiologic analysis: liver fat levels are a prognostic biomarker for incident liver outcomes

In the UK Biobank, we estimated the association between baseline liver fat and risk of incident liver cirrhosis, hepatocellular carcinoma, liver transplant or liver-related death. In a Cox proportional hazards regression model adjusting for age and sex, each 5% absolute increase PDFF was associated with a 64% increased risk of liver outcomes (hazard ratio = 1.64; 95% CI: 1.38-1.95; P = 2.4×10^-8^). The incidence in people with liver fat below 5% (a cutoff currently used in clinical practice to define the absence of hepatic steatosis at MRI imaging) was 0.035 per 1,000 person-years (7 events per 49,826 person-years of follow-up). Compared to having liver fat below 5%, having liver fat 10% was associated with 8.1-fold higher hazard of incident liver outcomes (**Figure 3**). These data indicate that, in the general population, the levels of baseline liver fat are strongly associated with the risk of incident cirrhosis with a large-effect association – whereby people without steatosis have very low absolute risk of cirrhosis.

**Figure 3:**
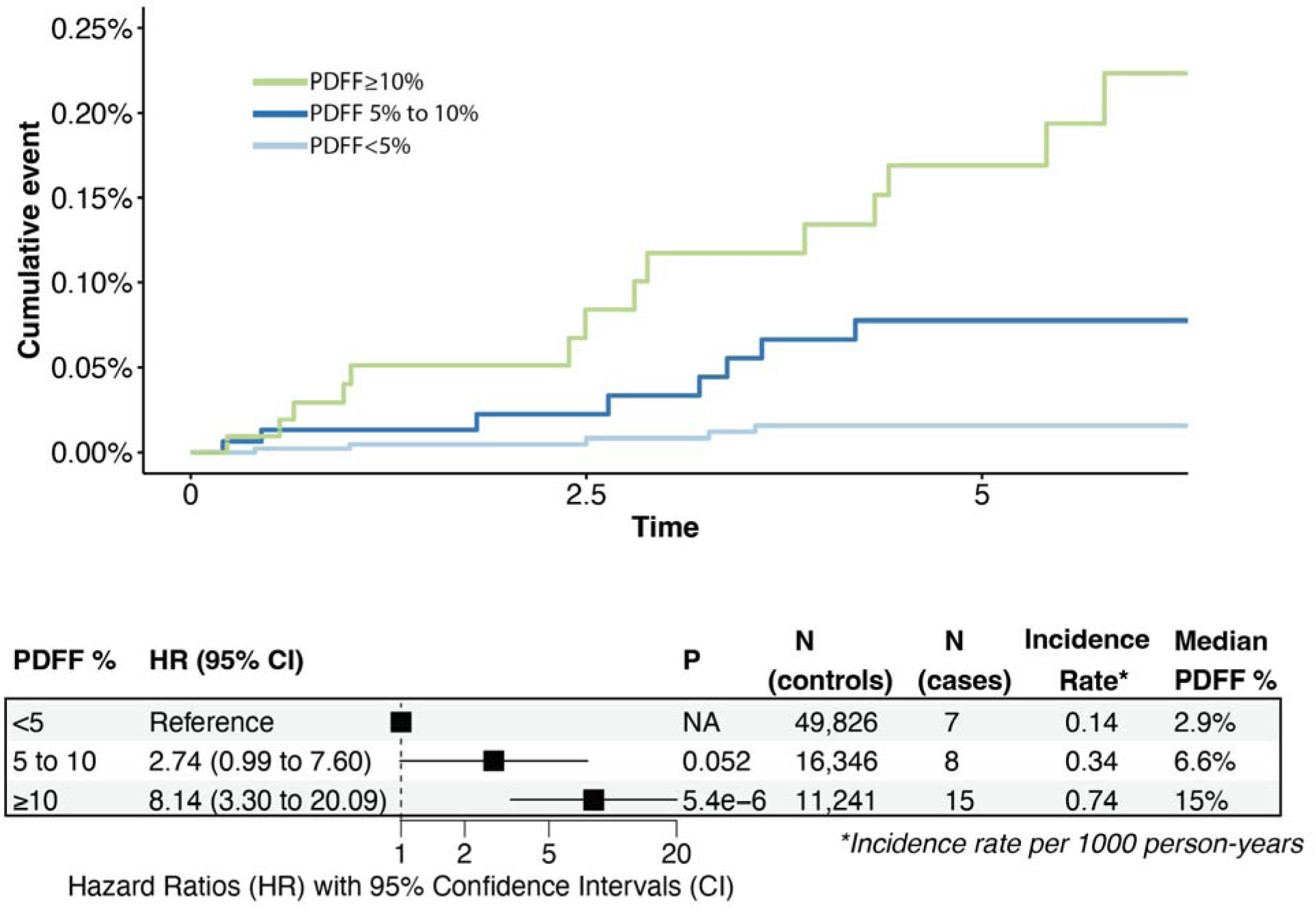
Increased Liver Fat by PDFF is a Strong Predictor for New-onset Liver Cirrhosis, Hepatocellular Carcinoma, Liver Transplant or Primary Liver Related Deaths in the UK Biobank.

### Meta-analysis of randomized controlled trials: liver fat reduction predicts histological improvement in MASH

We next performed a systematic review and meta-analysis of randomized clinical trials in MASH. The systematic review identified 20 phase 2 or 3 randomized controlled trials of investigational therapies for MASH that met the inclusion criteria (**Table 1, Supplementary Table 5, Supplementary Figure 4**). All included trials were prospective, randomized, placebo-controlled, blinded phase 2 or 3 studies with centralized, masked assessment of PDFF and histological endpoints using standardized protocols. All trials were prospectively registered on ClinicalTrials.gov with pre-specified endpoints, and the inclusion of multiple neutral and negative trials across diverse mechanisms reduces concern for publication bias.

**Table 1.** **Aggregate baseline summary, more details are in supplementary table 5 and 6** The overall means are weighted by the number of patients enrolled in each arm, giving more weight to larger arms. The ranges represent variation across arms: the minimum and maximum values observed across individual treatment arms. Footnote: †MASH resolution: ballooning = 0, inflammation ≤ 1, no fibrosis worsening. ‡Fibrosis improvement: ≥1-stage fibrosis reduction without MASH worsening.

| Characteristic, unit |  | Notes |
| --- | --- | --- |
| Total Studies, N | 20 | Phase 2/3 RCTs included in meta-analysis |
| Total Patients Enrolled, N | 4,259 | Across all arms |
| Total Patients Analyzed (MASH resolution endpoint†), N | 3,502 | 69 Arms with complete MASH + PDFF data |
| Total Patients Analyzed (Fibrosis improvement endpoint‡) | 3,502 | 69 Arms with complete Fibrosis + PDFF data |
| Study Duration, median (range) in weeks | 48 (12-52) |  |
| Treatment Arms, N (N patients) | 50 (2,534) | Active treatment arms analyzed |
| Placebo Arms, N (N patients) | 19 (968) | Placebo arms analyzed |
| **mean (min-max)** |  |  |
| Age, in years | 55 (47-60) | Weighted mean (range across arms) |
| BMI, kg/m<sup>2</sup> | 36 (31-39) | Weighted mean across 53 arms with data |
| Female, % | 59 (0-78) | Weighted mean across 64 arms with data |
| Type 2 Diabetes, % | 57 (32-86) | Weighted mean across 56 arms with data |
| Baseline PDFF, % | 19.1 (16-30) | Weighted mean across 64 arms with data |
| Baseline ALT, U/L | 59.2 (33-76) | Weighted mean across 48 arms with data |

The included studies enrolled a total of 3,502 patients with biopsy-confirmed MASH and reported both liver fat changes and histological endpoints; enrolled = 4,363 (**Supplementary Table 6, Supplementary Notes**). Study duration ranged from 20 to 52 weeks (median 48 weeks), with 50 treatment arms and 19 placebo arms analyzed. The most common drug mechanisms included thyroid hormone receptor-β (THR-β) agonists (3 trials, 7 arms), FGF21 analogues (3 trials, 12 arms – 6 analyzed), DGAT2 inhibitors (2 trials, 9 arms – 8 analyzed), see **Supplementary Table 7** for full details.

To estimate the relationship between liver fat changes (expressed as % relative change from baseline) and histological outcomes, we fitted linear mixed-effects meta-regression models. Specifically, to maximize statistical power and the spread in values of liver fat changes, we used each trial arm as the observation unit and to account for the dependence of different arms in the same trial, we modelled the trial identity as a random-intercept clustering variable. This approach allows greater generalizability as it maximises the spread of liver fat change and outcome occurrence values, while controlling for within-trial dependence of observations. Across 69 treatment or placebo arms from 20 phase 2 or 3 trials, mixed-effects meta-regression demonstrated that greater reductions in liver fat measured by PDFF were significantly associated with higher rates of MASH resolution without fibrosis worsening (β = -0.70%; 95% CI, -0.84% to -0.57%; *P* < 0.001; **Figure 4A, Supplementary Table 8**). Here, β reflects the difference in incidence of the outcome at the end of the trial in percentage units per 1 percentage-point relative decrease in liver fat by PDFF between baseline and end of follow-up. So, for each additional 10 percentage-point relative decrease in liver fat from baseline to end of follow-up, there was a 7 percentage-point higher MASH resolution.

**Figure 4:**
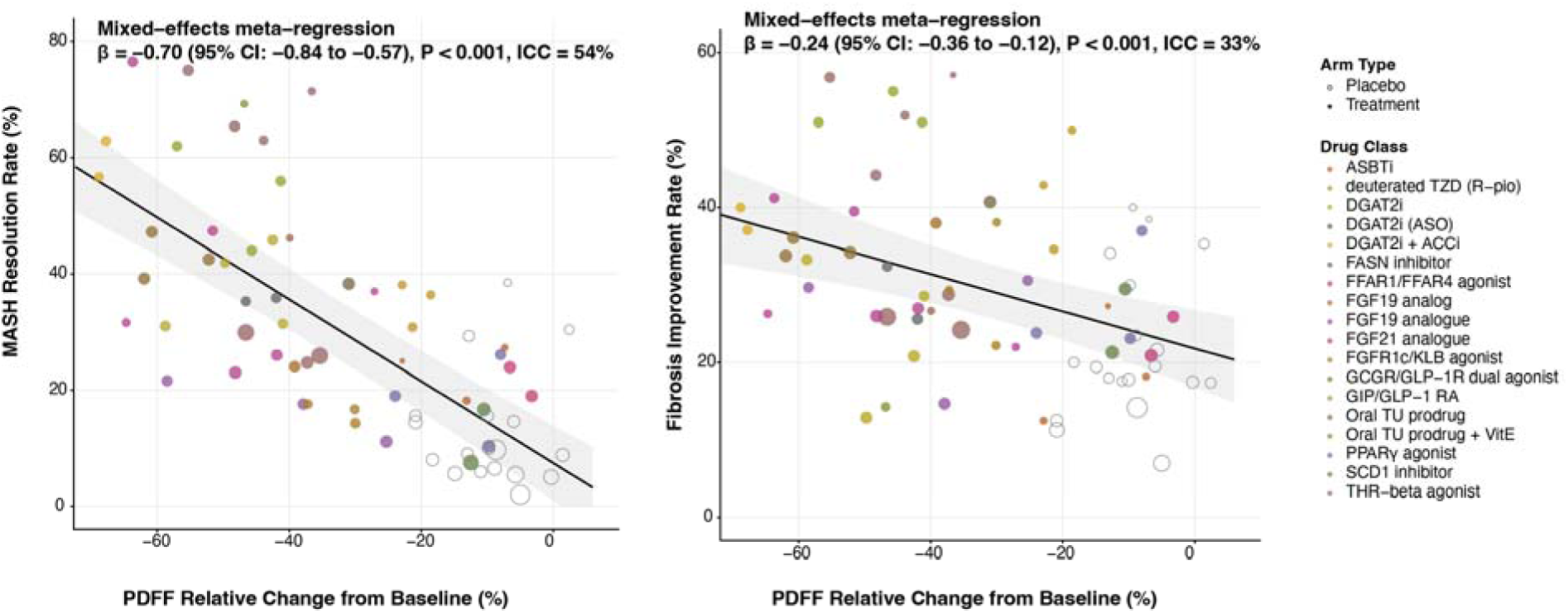
Trial-Level Analyses Demonstrate that the Reduction from Baseline in Liver Fat by PDFF is Strongly Associated with Histological Improvement. (A) Greater reductions in liver fat by PDFF are associated with higher rates of MASH resolution without fibrosis worsening (B) Greater reductions in liver fat by PDFF are associated with higher rates of fibrosis improvement without MASH worsening.

Liver fat reduction was also significantly associated with fibrosis improvement without MASH worsening, though the association was less pronounced (β = -0.24%; 95% CI, -0.36% to -0.12%; *P* < 0.001; **Figure 4B**). So, for each additional 10 percentage point relative decrease in liver fat from baseline to end of follow-up, there was a 2.4 percentage point higher fibrosis improvement.

In a secondary descriptive analysis stratifying study arms by quartiles of relative PDFF reduction, greater liver fat reduction was associated with progressively higher pooled NASH resolution rates. Pooled NASH resolution was 43.7% in the quartile with the greatest PDFF reduction, 32.8% in the second quartile, 18.0% in the third quartile, and 13.0% in the quartile with the smallest PDFF reduction. A similar but less pronounced pattern was observed for fibrosis improvement, with pooled rates of 34.0%, 32.2%, 23.8%, and 22.1% across quartiles. Restricted cubic spline analyses did not provide evidence of substantial non-linearity, supporting an overall monotonic association (**Supplementary Figure 5**).

Sensitivity analyses confirmed the robustness of the main analyses addressing potential sources of bias and demonstrating methodological robustness (**Supplementary Tables 9, 10 and 11**). Leave-one-out analysis demonstrated that no single study had a disproportionate influence (MASH β range, -0.73% to -0.66%; fibrosis β range, -0.29% to -0.21%; **Supplementary Table 11**). Covariate-adjusted meta-regressions and mixed-effects meta-regressions evaluated whether confounding by study characteristics could explain the PDFF-histology relationship; the PDFF coefficient for MASH resolution remained significant after adjustment for study duration, baseline PDFF level, or arm type (**Supplementary Table 9**), indicating that the association was independent of these design features.

## Discussion

Large human genetics and clinical trial data from this study demonstrate the strong etiologic relationship between liver fat and clinical outcomes in MASH.

Our study substantially expands the literature on this topic in many respects. First, we performed the largest liver fat GWAS including ∼80,000 people with liver MRI. We identified 56 (31 novel) loci for liver fat, more than doubling known liver fat associated genomic signals. This allowed us to develop a robust PRS for liver fat and estimate associations with cirrhosis and other severe liver outcomes in an independent set of >700,000 people. With this large scale analysis we demonstrate a strong association between liver fat loci and cirrhosis risk, with extreme consistency and a clear dose-response relationship both at the genomic locus (i.e. mechanism) level and at an individual-participant level, as demonstrated in the quintile individual-level analysis. While prior genetic studies have established that liver fat-associated loci associate with cirrhosis ^14-16,20-22^, our study quantifies this relationship in clinically actionable units, a 30% relative reduction in liver fat corresponds to a 39% lower odds of cirrhosis, and benchmarks its magnitude against a validated surrogate endpoint paradigm, i.e. the relationship between LDL cholesterol and coronary artery disease. Therefore, our genetic analysis shows that lifelong exposure to higher liver fat causes, independent of the mechanism driving it, a marked increase in cirrhosis risk, while lifelong exposure to lower liver fat is associated with protection from cirrhosis.

Separately, in a meta-analysis of 20 randomized controlled trials including >3,500 MASH patients, we show that the reduction in liver fat levels is a strong predictor of biopsy-based outcomes^10,23,24^, particularly MASH resolution. As in the genetic analysis, this relationship appears highly generalizable across mechanisms that induce a reduction in liver fat. Similar to genetic studies, also in this clinical trial meta-analysis there was a strong, continuous dose-response relationship without evidence of a cutoff for the observed effects. The implication is that, regardless of the mechanism, the reduction in liver fat attained by an intervention is likely translate into a benefit on liver bioposy endpoints. While in the clinical trial analysis we did not have access to individual level data, previous analyses of individual-level data from specific trials strongly complement and support our results. An individual participant data meta-analysis of seven early-phase trials comprising 346 participants demonstrated that a ≥30% relative decline in PDFF was associated with histological response and NASH resolution^24^. More recently, analyses of phase 2 MASH clinical trials have continued to use binary thresholds, demonstrating that ≥30% and ≥50% PDFF declines associate with MASH resolution and fibrosis regression at an individual patient level.^25^

The major implication of our study is that liver fat is a strong candidate as a reasonably likely surrogate endpoint for MASH clinical development. Notably, our data mirrors those for LDL-C in coronary disease, where LDL-C is a validated surrogate endpoint for coronary artery disease. For LDL-C, human genetics studies from the literature and our analysis show that lifelong LDL-C reductions due to genotype are associated with lower coronary disease risk with a magnitude of association similar to the one we observed in our data for liver fat and cirrhosis. Similarly, in clinical trials, LDL-C reductions predict the cardiovascular efficacy of LDL-C lowering treatments across a variety of mechanisms^26^.

The current paradigm for MASH clinical trials relies on liver biopsy, an invasive procedure with significant limitations. While biopsy has been the historical gold standard, it carries risks of morbidity and mortality, presents challenges for patient recruitment^27,28^, and is limited by sampling variability and low inter-pathologist concordance ^29^. In contrast, PDFF offers a non-invasive, reliable, and reproducible measure of liver fat with excellent repeatability, with a repeatability coefficient of 1.19% for same-scanner measurements on different days which matches clinical trial conditions.^30,31^ This technology provides comprehensive volumetric coverage of the entire liver, is widely available on modern MRI scanners, and has been incorporated into AASLD clinical guidelines for MASLD evaluation.^32^ The FDA has also recognized the challenges of tissue biopsies in clinical trials, encouraging alternative approaches for high-risk tissue sites, making PDFF an attractive alternative for MASH clinical development^33^.

Prior work examining the association between therapeutic reductions in liver fat and histological response has relied on binary PDFF thresholds in small early phase trials. Across genetic studies and clinical trial data, our results show that the etiologic relationship between liver fat and liver disease outcomes follows a continuous dose-response relationship, whereby larger reductions in liver fat for a longer duration of time should result in greater clinical benefit. Our study also shows the generalizability across mechanisms of the relationship between hepatic fat reduction and histological improvement.

## Conclusion

Large-scale human genetic studies and randomized-controlled trial data demonstrate that there is a strong, dose-dependent relationship between liver fat and liver disease severity, histological changes and clinical liver events in the general population and in MASH patients. These data support the use of liver fat reduction as a reasonably likely surrogate endpoint for MASH clinical development.

### Online Methods

We conducted three sets of analyses. First, using genotype sequencing data, we conducted a ∼700,000-participant two sample mendelian randomization (MR) analysis, an approach that uses genetic variants associated with a certain exposure or biomarker (in this case, liver fat percentage by PDFF) as instrumental variables to understand the causal relationship of the exposure with a disease outcome (liver cirrhosis, hepatocellular carcinoma and fibrosis grade). Second, we analyzed the association of liver fat assessed by PDFF with new clinical liver events using UK Biobank data. Finally, we performed a meta-analysis of the association between the change from baseline in liver fat assessed by PDFF and registrational histological endpoints in published MASH clinical trials.

### DNA sequencing data

The Regeneron Genetics Center (Tarrytown, NY, USA) performed high-coverage exome sequencing using Illumina v4 HiSeq 2500 or NovaSeq instruments, with 75 base pairs paired-end reads. The GRCh38 Human Genome reference sequence and Ensembl v100 gene definitions were used for variant identification and annotation.

### Common variant genotyping and imputation

For common variant association analyses, we used genotyping data generated by UKB as described.^34^ In the remaining cohorts, common variant genotyping was performed using Illumina Human Omni Express Exome and Global Screening arrays.^35^ Genome-wide imputation was performed using the TOPMed reference panel^36^.

### Participating Cohorts

Genotyping data was available for up to 1,049,842 individuals across 10 multi-ancestry cohorts, though sample sizes vary by phenotype:

1. 488,201 from the population-based UK Biobank (UKB),^37^
2. 169,467 from the Geisinger Health System MyCode cohort (GHS),^38^
3. 116,226 from the health system based Mayo Clinic Project Generation (MAYO-RGC) which includes the Mayo Clinic Biobank and 30 disease registries,^39^
4. 72,710 from the health system based Colorado Center for Personalized Medicine (CCPM) Biobank,^40^
5. 63,897 from the health system based UCLA ATLAS Precision Health Biobank (ATLAS),^41^
6. 57,898 from the health system based Mount Sinai BioMe Biobank (BioMe),
7. 41,971 from the health system based University of Pennsylvania Penn Medicine Biobank (PMBB),^42^,
8. 30,104 from the Malmö Diet and Cancer Study (MDCS).^43^
9. 6,010 from the Indiana University School of Medicine (Indiana-CLDB) study
10. 3,358 from the NASH-CRN study^44^.

Each study was approved by their respective institutional review boards, with appropriate informed consent procedures followed according to local regulations and guidelines.

### Phenotype definitions

#### Magnetic resonance imaging (MRI) of the liver in UKB

A subset of participants in UKB underwent magnetic resonance imaging (MRI) of the liver.^45^ Liver fat quantification was performed using two distinct acquisition protocols: an initial cohort of ∼10,000 participants underwent Dixon gradient-echo imaging, while subsequent participants (imaged from 2016 onward) were scanned using an IDEAL (Iterative Decomposition of water and fat with Echo Asymmetry and Least-squares estimation) sequence. All acquisitions utilized Siemens MAGNETOM clinical scanners with standardized parameters: 2.5×2.5 mm in-plane resolution and 6 mm slice thickness, yielding complex-valued 2D datasets per participant.

Measurements of hepatic fat percentage (proton density liver fat fraction; PDFF), a measure of the proportion of fat content in the liver, were obtained by applying pre-defined mathematical models after segmenting the liver on liver MRI images.^46^ The implementation was validated using a publicly available phantom dataset containing vials of varying concentrations of fat.^47^ We estimated PDFF as the fraction of fat signal relative to total fat plus water signal. Pixels belonging to the liver were segmented using a Li thresholding approach for PDFF maps to identify liver tissue. To obtain a summary measure of each trait per subject, all pixels within the liver were averaged for each parametric map. PDFF was then used as the outcome for the analysis. For further details on the derivation of these phenotypes please see O’dushlaine et al.^48^ and Verweij et al.^49^ . A separate dataset of 10,377 individuals that was released as part of 2026 imaging data release, was analyzed separately from the GWAS discovery.

### Liver disease phenotypes used in the Mendelian randomization and polygenic risk score analysis

We defined cases of binary liver disease outcomes based on one or more of the following criteria: (i) self-reported disease obtained from digital questionnaire or interview with a trained nurse, (ii) in-patient hospitalization for the disease or clinical-problem list entries of the disease according to International Classification of Diseases, Ninth (ICD-9) or Tenth (ICD-10) Revision diagnosis code, (iii) medical procedures or surgery due to the disease, (iv) death due to the disease, and (v) a disease diagnosis code entered for two or more outpatient visits in separate calendar days. In short, liver cirrhosis was defined by K703(Cirrhosis), K704(Alcoholic hepatic failure), K717(Toxic liver disease with fibrosis and cirrhosis of liver), K721(Chronic hepatic failure), K746(Other and unspecified cirrhosis of liver); hepatocellular carcinoma C220(Liver cell carcinoma). We defined controls as individuals who did not meet any of criteria for case status. To minimize misclassification, we excluded from the control group (i) non-cases diagnosed with any type of liver disease (not restricted to the type of liver disease in question), (ii) non-cases with only one out-patient encounter related to the type of liver disease in question, (iii) non-cases diagnosed with ascites presumably related to liver failure, and (iv) non-cases with elevated alanine aminotransferase (ALT) levels (>33 IU/L for men^50^, >25 IU/L for women^50^), please see Verweij et al.^49^ for more details.

### Genome Wide Association Study of Liver Fat and Statistical Genetics Analyses

We performed a genome-wide association study of liver fat percentage (PDFF-MRI) adjusted for body mass index using REGENIE v3.4. Prior to analysis, liver fat values were rank inverse normal transformed (RINT) separately for each sex to ensure normality.

Genetic associations were tested using linear regression models implemented in REGENIE to account for relatedness and population structure. Analyses were adjusted for age, age², sex, age-by-sex and age²-by-sex interaction terms, experimental batch-related covariates, the first 10 common variant-derived principal components (PCs), and a polygenic score generated by REGENIE that controls for relatedness and population structure.

All genetic analyses were adjusted for age, age^2^, sex, age-by-sex and age^2^-by-sex interaction terms, experimental batch-related covariates, principal components and accounted for a polygenic score generated by REGENIE that controls for relatedness and population structure.^51^. Results were combined across subsets by fixed-effect inverse variance-weighted meta-analysis.

### Polygenic risk score analyses and Mendelian Randomization

We estimated the causal relationship between liver fat percentage (PDFF measured by MRI as described above) and liver disease outcomes using a two-sample Mendelian randomization (MR) or polygenic risk score framework. Genetic instruments were taken from the genome-wide association study of liver fat percentage (PDFF-MRI) adjusted for body mass index. From the top hits, we performed LD clumping using PLINK with parameters: MAF > 0.01, r² < 0.001, P < 5×10^-8^. This yielded 56 independent genetic variants. Variants in the HFE gene region were excluded to avoid potential pleiotropic effects through iron metabolism pathways, resulting in a final set of genetic instruments. For the exposure (liver fat percentage), we used effect estimates from an unadjusted PDFF GWAS. For outcomes, we re-estimated variant-outcome associations excluding samples that contributed to the PDFF discovery GWAS to ensure independence between exposure and outcome datasets. We meta-analyzed results across cohorts where applicable.

Two-sample MR were performed using multiple methods to assess robustness analyses including two inverse-variance-weighted MR models^52^; weighted median MR^52^; and MR-Egger^53^. Analyses were conducted using the TwoSampleMR R package^54^. Genetic effects were converted to clinically meaningful units by expressing PDFF changes as percentage units relative to the population mean (5.8%). We calculated the effect of a 30% relative decrease in PDFF (corresponding to a 1.75% absolute decrease) and a 1 standard deviation decrease (5.4% absolute decrease) on disease odds. The results demonstrated that genetically predicted lower liver fat was causally associated with reduced odds of liver disease outcomes, with consistent effects across multiple MR methods.

### Survival analysis in UK Biobank

The primary composite outcome comprised new-onset liver cirrhosis, hepatocellular carcinoma, liver transplantation, or liver-related death. Liver cirrhosis was defined using ICD-10 codes K703 (alcoholic cirrhosis), K704 (alcoholic hepatic failure), K717 (toxic liver disease with fibrosis and cirrhosis), K721 (chronic hepatic failure), and K746 (other and unspecified cirrhosis). Hepatocellular carcinoma was identified using ICD-10 code C220 (liver cell carcinoma). Liver transplantation was ascertained through OPCS-4 procedure codes, and liver-related death was defined as any primary cause of death with liver disease ICD-10 codes (K70-K77, I81, I85, I982, I983, I864, T864, Z944, C220). Prior liver cirrhosis was also established by nurse-interview at the UK Biobank visits (liver failure cirrhosis field 1158 and alcoholic liver disease alcoholic cirrhosis field 1604).

Follow-up time was calculated from the imaging visit date to the earliest occurrence of any outcome event, death from other causes, or administrative censoring. Censoring dates corresponded to complete mortality data availability: 31 May 2024 for England and Wales, or 31 December 2023 for Scotland. Participants with negative follow-up time or historical evidence of cirrhosis or hepatocellular carcinoma were excluded from analyses.

We performed Cox proportional hazards regression to estimate hazard ratios (HRs) and 95% confidence intervals (CIs) for liver fat categories (<5%, 5-10%, ≥10% PDFF) relative to the reference category (<5% PDFF). Models were adjusted for age at imaging visit and sex. The proportional hazards assumption was assessed using Schoenfeld residuals. Kaplan-Meier curves were generated to visualize cumulative incidence by liver fat category. We additionally examined continuous associations between liver fat percentage and liver outcomes.

All survival analyses were conducted using R version 4.3.0 with the survival and survminer packages. Two-sided P-values <0.05 were considered statistically significant.

### Liver histopathologic phenotype definitions

In the Geisinger Health System bariatric surgery cohort, we analyzed liver biopsies from 3,599 individuals of European descent who underwent bariatric surgery and were enrolled in the GHS MyCode and GHS-Regeneron Genetics Center DiscovEHR collaboration^38^. During surgery, wedge liver biopsies were obtained 10 cm to the left of the falciform ligament prior to liver retraction or gastric procedures, following a standardized protocol. Biopsy specimens were divided into sections: the primary section was processed for clinical histopathology (fixed in 10% neutral buffered formalin, stained with hematoxylin and eosin for routine histology, and Masson’s trichrome for fibrosis assessment), while remaining sections were preserved in a research biobank using RNAlater tissue collection system (ThermoFisher Scientific) or liquid nitrogen storage.

An experienced hepatopathologist performed initial histological examinations, with subsequent independent review by a second pathologist. All specimens were scored according to the NASH Clinical Research Network histological scoring system^55^.

In the NASH Clinical Research Network cohort, we conducted a cross-sectional analysis of participants enrolled in the Nonalcoholic Steatohepatitis Clinical Research Network (NASH-CRN), a prospective multicenter cohort recruiting patients with biopsy-confirmed metabolic dysfunction-associated steatotic liver disease (MASLD) across the United States from 2004 to 2020. All participants provided written informed consent, and the study protocol conformed to the ethical guidelines of the 1975 Declaration of Helsinki.

All liver biopsies underwent blinded review by the NASH-CRN Pathology Committee using the standardized NASH-CRN scoring system. Fibrosis was staged from 0 (no fibrosis) to 4 (cirrhosis), steatosis graded 0-3, hepatocyte ballooning 0-2, and lobular inflammation 0-3. The NAFLD Activity Score was calculated as the sum of steatosis, ballooning, and inflammation scores.

The NASH-CRN study was approved by the NASH-CRN Steering Committee and followed NIH Genomic Data Sharing policies.

### Systematic Meta-analysis

This systematic review and meta-analysis was conducted and reported in accordance with the Preferred Reporting Items for Systematic Reviews and Meta-Analyses (PRISMA) 2020 guidelines (**Supplementary Figure 2**). The duplicate removal and screening of titles and abstracts were performed independently by two reviewers. Discrepancies were resolved by consensus, or by a third reviewer. Full-text assessment of potentially eligible studies was also conducted in duplicate by the same two reviewers.

This review protocol was registered with the International Prospective Register of Systematic Reviews (PROSPERO) database (CRD42025649746; available from: https://www.crd.york.ac.uk/PROSPERO/view/CRD42025649746). The meta-analysis was conducted as part of a broader investigation combining genetic, observational, and trial-level evidence to evaluate liver fat as a surrogate endpoint for MASH.

We systematically searched PubMed, Embase, and ClinicalTrials.gov for randomized clinical trials of metabolic dysfunction-associated steatohepatitis (MASH) that reported both MRI-derived proton density fat fraction (PDFF) and histological outcomes. Studies were required to report PDFF changes from baseline and at least one histological endpoint (MASH resolution or fibrosis improvement by ≥1 stage). We extracted data at the treatment arm level, including both active treatment and placebo arms.

#### Eligibility criteria

Studies were included if they met all of the following criteria: (1) randomized clinical trial for non-alcoholic steatohepatitis (NASH) or metabolic dysfunction-associated steatohepatitis (MASH), at phase 2 or 3; (2) enrolled adults aged 18 years or older; (3) participants had biopsy-confirmed MASH with fibrosis stages F1–F3 and NAFLD Activity Score (NAS) >=4; (4) reported arm-level summary statistics for percent change from baseline in liver fat measured by PDFF (or, if percent change was unavailable, both baseline and change-from-baseline values, or baseline and endpoint values sufficient to calculate relative change), together with at least one of the following histological endpoints: proportion of participants achieving MASH resolution without fibrosis worsening, or proportion achieving fibrosis improvement (>=1 stage) without MASH worsening; and (5) the study was reported in a peer-reviewed article, conference abstract, or press release.

MASH resolution is defined as achieving all three of the following criteria on end-of-treatment liver biopsy: (1) hepatocyte ballooning score = 0, (2) lobular inflammation score ≤ 1, and (3) no worsening of fibrosis (defined as no increase in fibrosis stage ≥ 1 point) compared to baseline biopsy. This composite endpoint represents complete resolution of steatohepatitis activity while ensuring liver fibrosis did not progress during treatment.

Fibrosis improvement is defined as a decrease in fibrosis stage ≥ 1 point on the NASH-CRN fibrosis scoring system (range 0-4) from baseline to end-of-treatment biopsy, without worsening of MASH (defined as any increase in either hepatocyte ballooning score or lobular inflammation score). This ensures that fibrosis improvement is not achieved at the expense of worsening steatohepatitis activity.

Studies were excluded if any of the following applied: phase 1, observational, or phase 4 design; enrolment of pediatric or adolescent participants (age <18 years); inclusion of participants with F4 fibrosis, cirrhosis, or hepatocellular carcinoma at baseline; non-English language publication; or results not available through peer-reviewed journals, conference abstracts, or press releases.

#### Database searches

##### All database searches were conducted on November 17, 2025

**Embase** (150 records). The search was constructed in five steps: (1) disease concept – ‘non-alcoholic steatohepatitis’/exp OR ‘non-alcoholic steatohepatitis’ OR (‘non alcoholic’ AND (‘steatohepatitis’/exp OR steatohepatitis)) OR ‘metabolic-associated steatohepatitis’ OR (‘metabolic associated’ AND (‘steatohepatitis’/exp OR steatohepatitis)) (26,959 records); (2) histological confirmation – ‘liver biopsy’ (89,952 records); (3) combined disease and diagnostic method – #1 AND #2 (5,615 records); (4) study design filter – #3 AND ‘randomized controlled trial’/de (351 records); (5) study phase filter – #4 AND (‘phase 2 clinical trial’/de OR ‘phase 3 clinical trial’/de) (150 records).

**PubMed MEDLINE** (190 records). The search combined three concepts with AND operators: (1) disease/condition – “non alcoholic fatty liver disease”[MeSH Terms] OR (“non alcoholic”[All Fields] AND “fatty”[All Fields] AND “liver”[All Fields] AND “disease”[All Fields]) OR “non alcoholic fatty liver disease”[All Fields] OR (“non”[All Fields] AND “alcoholic”[All Fields] AND “steatohepatitis”[All Fields]) OR “non alcoholic steatohepatitis”[All Fields] OR (“Metabolic-Associated”[All Fields] AND (“fatty liver”[MeSH Terms] OR (“fatty”[All Fields] AND “liver”[All Fields]) OR “fatty liver”[All Fields] OR “steatohepatitis”[All Fields])); (2) diagnostic method – ((“liver”[MeSH Terms] OR “liver”[All Fields] OR “livers”[All Fields] OR “liver s”[All Fields]) AND “biopsy”[MeSH Terms]) OR “MASH”[All Fields]; (3) study design – randomizedcontrolledtrial[Filter].

**ClinicalTrials.gov** (127 records). The search was constructed in five steps: (1) disease concept – AREA[BasicSearch] (Non-Alcoholic Steatohepatitis OR Metabolic-Associated Steatohepatitis) (2,158 records); (2) histological confirmation – AND (Liver Biopsy) (457 records); (3) age restriction – AND AREA<u>StdAge</u> (446 records); (4) study phase – AND AREA<u>Phase</u> (127 records); (5) study type – AND AREA<u>StudyType</u> (127 records).

To ensure comprehensive capture of recently completed trials, we employed multiple supplementary search strategies: 1. Systematic hand-searching of conference proceedings from AASLD (The Liver Meeting). 2. Manual review of ClinicalTrials.gov for all Phase 2/3 NASH trials marked as ‘Completed’ to identify studies with outcome data not yet published in peer-reviewed journals 3. For trials identified through these methods, we searched for outcome data through: Conference abstract databases, Pharmaceutical company press releases, PubMed (by NCT identifier), ClinicalTrials.gov results database. This approach identified 3 trials of which 2 were already included via the pubmed or ClinicalTrials.gov search, NCT04173065 (VOYAGE): AASLD 2024 abstract, PMID 39896964; NCT03551522 (Seladelpar): CT.gov results + press release; NCT04134091 (LiFT): CT.gov results + press release.

The complete study selection process, including the number of records identified, screened, and excluded at each stage, is presented in the PRISMA flow diagram (**Supplementary Figure 2**).

#### Data extraction

Data were extracted at the treatment-arm level, including both active treatment and placebo arms. For each arm, we recorded the mean relative change in PDFF from baseline (%), the rate of MASH resolution (%), the rate of fibrosis improvement (%), the number of patients analyzed for each endpoint, study duration, baseline patient characteristics (age, sex, BMI, prevalence of type 2 diabetes, baseline PDFF and baseline ALT), drug class, and whether the arm was placebo or active treatment arm.

#### Outcome and sampling variance

For each arm, the outcome was the proportion of patients achieving the histological endpoint, expressed as a percentage, as reported by the trial. Sampling variances were derived from the binomial distribution and rescaled to the percentage scale:

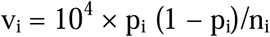

where p_i_ is the proportion of patients with the event and n_i_ is the number of patients analysed in arm i. Rescaling by 10^4^ (since Var(100p) = 10^4^ x Var(p)) places the sampling variances on the same scale as the between-arm variance (τ^2^). Without it, sampling variances would be negligible relative to τ^2^, and arms would be weighted almost equally regardless of sample size.

#### Primary Analysis

To account for the hierarchical structure of the data (multiple arms nested within trials), we fitted linear mixed-effects meta-regression models as the primary analysis. Modelling trial identity (NCT) as a random intercept controls for within-trial dependence of arms from the same study. For each outcome the model took the form:

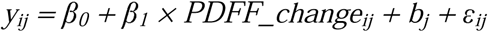

where *y*_ij_ is the observed outcome rate (%) for arm *i* in study *j*, *b*_j_ ∼ N(0, τ^2^) is the random study intercept capturing between-study variance, and ε_ij_∼ N(0, σ^2^) is the residual within-study error. Models were fitted using the lme4 package with *P* values and 95% confidence intervals obtained via lmerTest. The regression coefficient β_1_ quantifies the expected change in outcome rate (percentage points) per 1-percentage-point relative change in PDFF. The intraclass correlation coefficient ICC = τ^2^/(τ^2^ + σ^2^) measures the proportion of total variance attributable to between-study differences.

As a complementary approach, a three-level inverse-variance weighted meta-regression was fitted with the metafor package, so that arms are weighted by their precision while the clustering of arms within trials is retained:

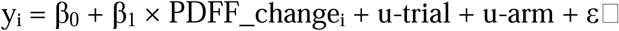

where ε_i_ ∼ N(0, v_i_) is within-arm sampling error with known variance v_i_ as defined above, and u-trial and u-arm are random effects at the trial and arm levels, specified as random = ∼ 1|NCT/arm. Variance components were estimated by restricted maximum likelihood (REML). This specification was chosen because a two-level model with an arm-level random effect alone ignores the nesting of arms within trials, while the unweighted mixed model ignores arm size; the three-level model addresses both. R^2^ was calculated as the proportion of between-arm heterogeneity explained by PDFF change, comparing the total random-effects variance from models with and without the moderator:

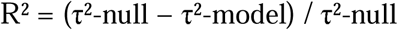

where τ² is the sum of the trial- and arm-level variance components.

We also performed a secondary arm-level analysis in which study arms were grouped into quartiles of relative PDFF change from baseline. Within each quartile, pooled NASH resolution and fibrosis improvement rates were estimated using random-effects meta-analysis of logit-transformed proportions, with back-transformation to percentages for presentation and a study-level random effect to account for within-trial clustering when multiple arms from the same trial contributed to the same model. These quartile-based analyses were descriptive and complementary to the primary continuous meta-regression.

#### Sensitivity Analyses

We performed pre-specified sensitivity analyses stratifying by: (1) treatment duration (≤24 weeks vs >24 weeks), (2) arm type (treatment vs placebo), (3) baseline PDFF severity (median split), and (4) analysis type (least-squares means vs arithmetic means), **Supplementary Table 9**. We conducted leave-one-out sensitivity analysis by sequentially excluding each study and re-fitting the model to assess whether any single study had disproportionate influence on the results (**Supplementary Table 11, Supplementary Figure 6**).

#### Heterogeneity Assessment

Between-arm heterogeneity was assessed using Cochran’s *Q* statistic and the *I*² statistic, estimated via REML-based random-effects modelling (metafor::rma). computed on the full analysis set of 69 arms so that the degrees of freedom are k − 1 for both outcome. *I*² values of 25%, 50%, and 75% were interpreted as low, moderate, and substantial heterogeneity, respectively. To partition the overall I² into between-study and within-study components, we used the multilevel I² approach, in which each variance component of the three-level model is expressed relative to the sum of the variance components and the typical within-arm sampling variance.

### Software

REGENIE v3.4.^12^ was used for the genome wide study. All analyses were performed in R (version 4.3.1). Meta-regression and heterogeneity statistics were computed using the metafor package (version 4.6). Mixed-effects models were fitted using lme4 (version 1.1) with P values obtained via the lmerTest package. Figures were generated using ggplot2.

## Data Availability Statement

All data needed to reproduce the manuscript’s analyses are available in **Supplementary Tables 2,5–7**, including all genetic variant statistics and details of randomized clinical trials (baseline characteristics, PDFF changes, and histological outcomes for the 69 treatment and placebo arms). Detailed notes documenting data sources, extraction methods, and assumptions for each study are provided in Supplementary Notes.

## Ethics approval and consent

All study procedures were conducted in accordance with the Declaration of Helsinki. Informed consent was obtained from all participants in each contributing cohort. More information is in the Supplement.

The UK Biobank received ethical approval from the North West Centre for Research Ethics Committee (11/NW/0382); this analysis was conducted under application number 26041. The Geisinger Health System MyCode Community Initiative was approved by the Geisinger IRB (Study #2006-0258; #2017-158). The Mayo Clinic Project Generation study was approved by the Mayo Clinic IRB (protocol #19-007763). The Colorado Center for Personalized Medicine Biobank was approved by the University of Colorado Anschutz Medical Campus IRB (protocol #15-0461). The UCLA ATLAS Precision Health Biobank was approved by the UCLA IRB (IRB#17-001013). The Mount Sinai BioMe Biobank and Million Health Discoveries Program were approved by the Icahn School of Medicine at Mount Sinai IRB (PPHS IRB #11-01139 and #21-01743). The Penn Medicine Biobank was approved under IRB protocol `#813913`. The Malmö Diet and Cancer Study received ethical approval from the Ethics Committee of Lund University (LU 51-90) and the Regional Board of Ethics in Lund (Dnr 2016/479). The Indiana University School of Medicine Indiana Biobank was approved by the Indiana University IRB (protocol #1105005445). The NASH CRN studies were approved at each participating center; oversight was provided by a data safety and monitoring board appointed by the NIDDK. Full details of ethics approvals for each cohort are provided in the Supplementary Information.

Green points represent genetic variants (55 PDFF-associated variants excluding HFE locus in A,C; 76 LDL-associated variants in B). Dashed lines show inverse-variance weighted two-sample Mendelian randomization estimates with 95% confidence intervals (shaded areas). In A and C, the effect of PNPLA3 p.I148M heterozygous and homozygous genotypes on PDFF and cirrhosis was added to the plot. In B the PCSK9 R49L allele and PCSK9 rare pLoF (allele frequency <1%) variants based on 2,034 carriers among 789,796 individuals were added to the plot.

Increased Liver fat by PDFF is a strong predictor for a primary diagnosis of liver cirrhosis (N=4), hepatocellular carcinoma (N=12), liver transplants (N=5) and primary liver related deaths (N=9) in the UK Biobank. Individuals with any evidence (primary or secondary diagnosis) of cirrhosis, hepatocellular carcinoma or liver transplant before the imaging visit were excluded. Survival analyses were adjusted for age and sex among 77,443 individuals that had follow-up (death censoring) beyond the imaging visit; median follow-up was 4.6 years.

Shown are the relationships between change from baseline in liver fat by PDFF and histological endpoints across 69 treatment and placebo arms from 20 randomized phase 2 or phase 3 clinical trials. Studies included in the meta-analysis had data publicly released on ClinicalTrials.gov, published in peer-reviewed journals, conference abstracts, or press releases, and contained PDFF changes and histological responses. MASH trials with populations that were pediatric, adolescent, cirrhotic (F4), or had hepatocellular carcinoma were excluded.

Each point represents a separate treatment or placebo arm. The x-axis shows the mean percent (%) change from baseline in liver fat by PDFF (negative values indicate reduction). The y-axis shows the percentage of patients achieving the histological endpoint. Point size reflects the number of patients analyzed (larger points = larger sample size). Point color indicates the drug class (therapeutic mechanism). Filled circles represent active treatment arms; open circles represent placebo arms. The solid line is the mixed-effects meta-regression fit (fixed effect from a linear mixed model with random intercept per study) with the shaded area showing the 95% confidence band. ICC (Intraclass Correlation Coefficient) represents proportion of total variance that is attributable to differences between studies.

