## Supplementary Information for "Causal relationship between liver fat reduction and clinical outcomes in human genetic studies and randomized clinical trials of metabolic-dysfunction associated steatohepatitis"

**Supplement**

### Supplementary Figures

#### Supplementary Figure 1

Quantile–quantile (Q–Q) plot of observed versus expected −log₁₀(P) values from the genome-wide association study (GWAS), restricted to 1,087,458 HapMap3 SNPs in 5,000 random individuals from the UK Biobank. The genomic inflation factor was λ_GC = 1.108. LD score regression (LDSC), fit using robust regression (rlm), estimated SNP-based heritability on the observed scale at h² = 0.229 (LD score-weighted regression of χ² statistics against LD score), with an intercept of 0.975. Because the intercept is close to 1 (attenuation ratio = (intercept − 1)/(mean χ² − 1) ≈ −0.02), the modest inflation in test statistics is attributable almost entirely to polygenic signal rather than to residual population stratification or other confounding. The dotted red line indicates genome wide significant.

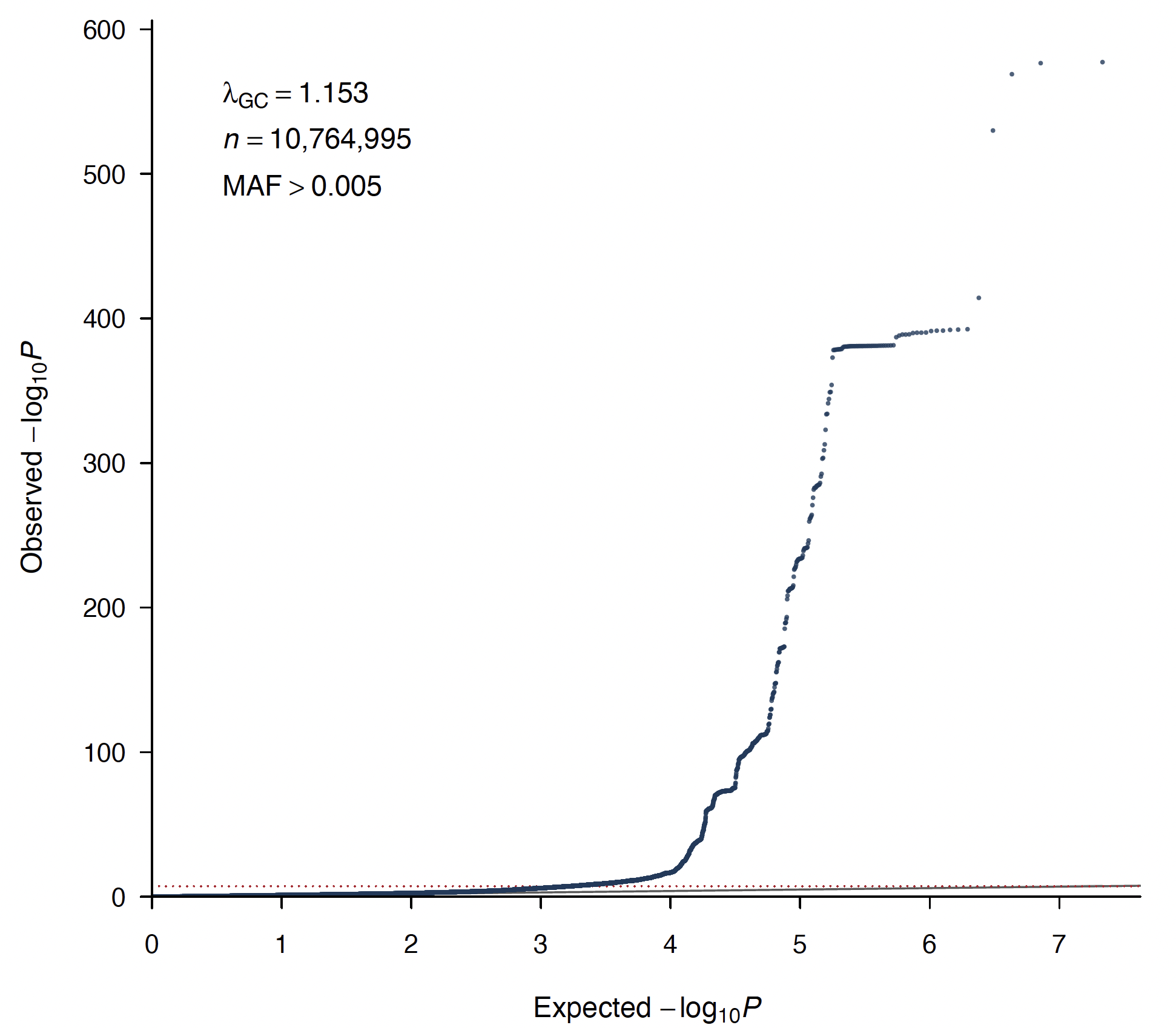

#### ****Supplementary Figure 2****

**Mendelian randomization (IVW-Fixed effects) leave-one-out sensitivity analysis for the effect of PDFF on liver cirrhosis (A) and hepatocellular carcinoma (B) per 30% lower PDFF.** Leave-one-out analyses of the IVW fixed-effects mendelian randomization estimate were performed to assess whether the IVW causal estimate of liver fat % by PDFF on liver disease outcomes was driven by any single genetic instrument. Each row shows the MR estimate (odds ratio, black point) and 95% confidence interval (horizontal line) obtained after excluding the labeled variant from the instrument set; "All" (red) shows the estimate using the full set of instruments. Estimates remained consistent regardless of which individual variant was excluded, indicating that the causal association between PDFF and liver disease outcomes was not driven by a single instrument in either genotype stratum. I148M (22:43928847:C:G) did contribute more to the overall effect, compared to the rest of the variants**,** but sensitivity analyses that are robust to outliers, including MR-Egger, weighted median, and weighted mode, are all consistent with a strong causal effect of liver fat on disease risk. (**Supplementary Table** **4**).

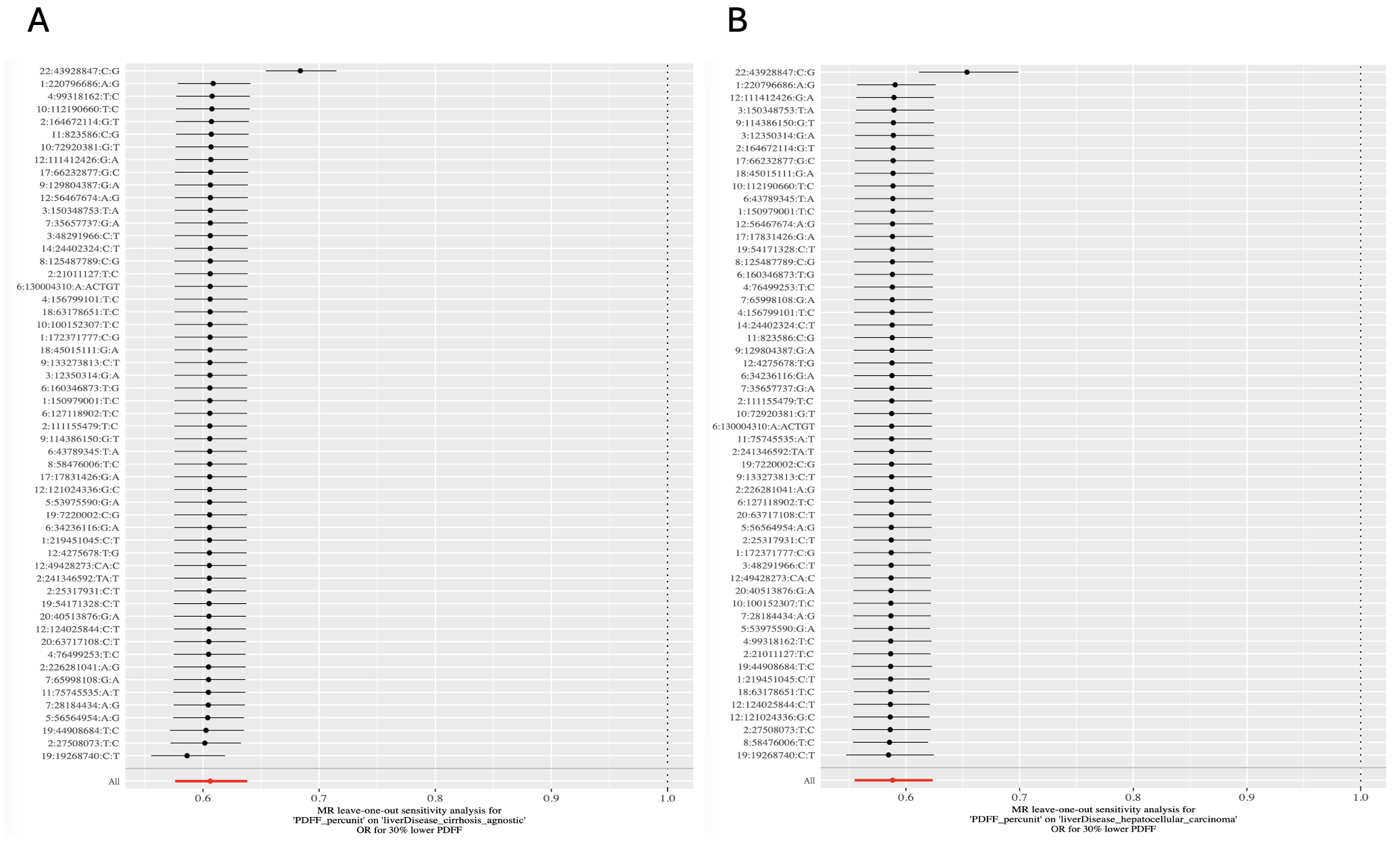

#### Supplementary Figure 3

A polygenic risk score (PRS) for PDFF is associated with Steatosis Score and Fibrosis Grade on Biopsy in the bariatric surgery cohort (A) and NASH-CRN Cohort (B), an extension of Figure 2.

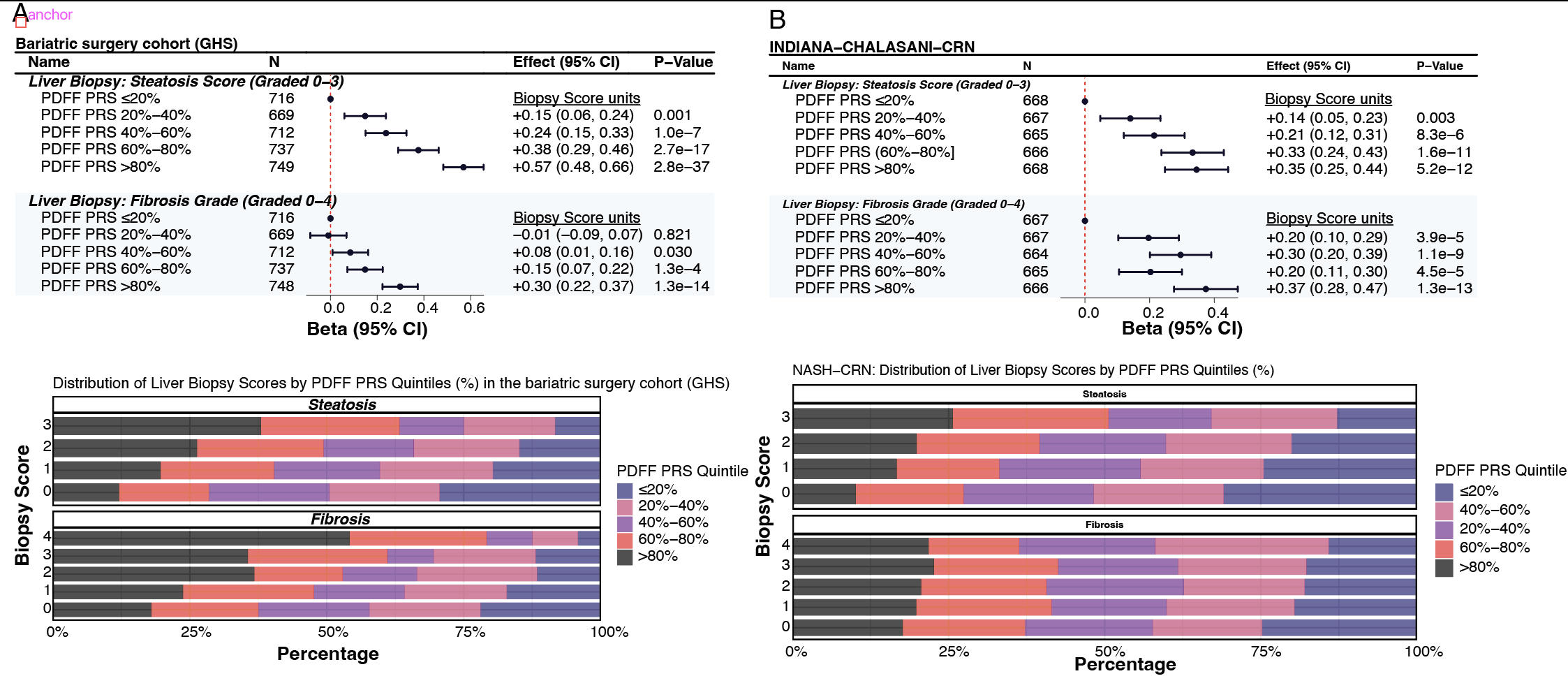

#### Supplementary Figure 4 Flow Diagram

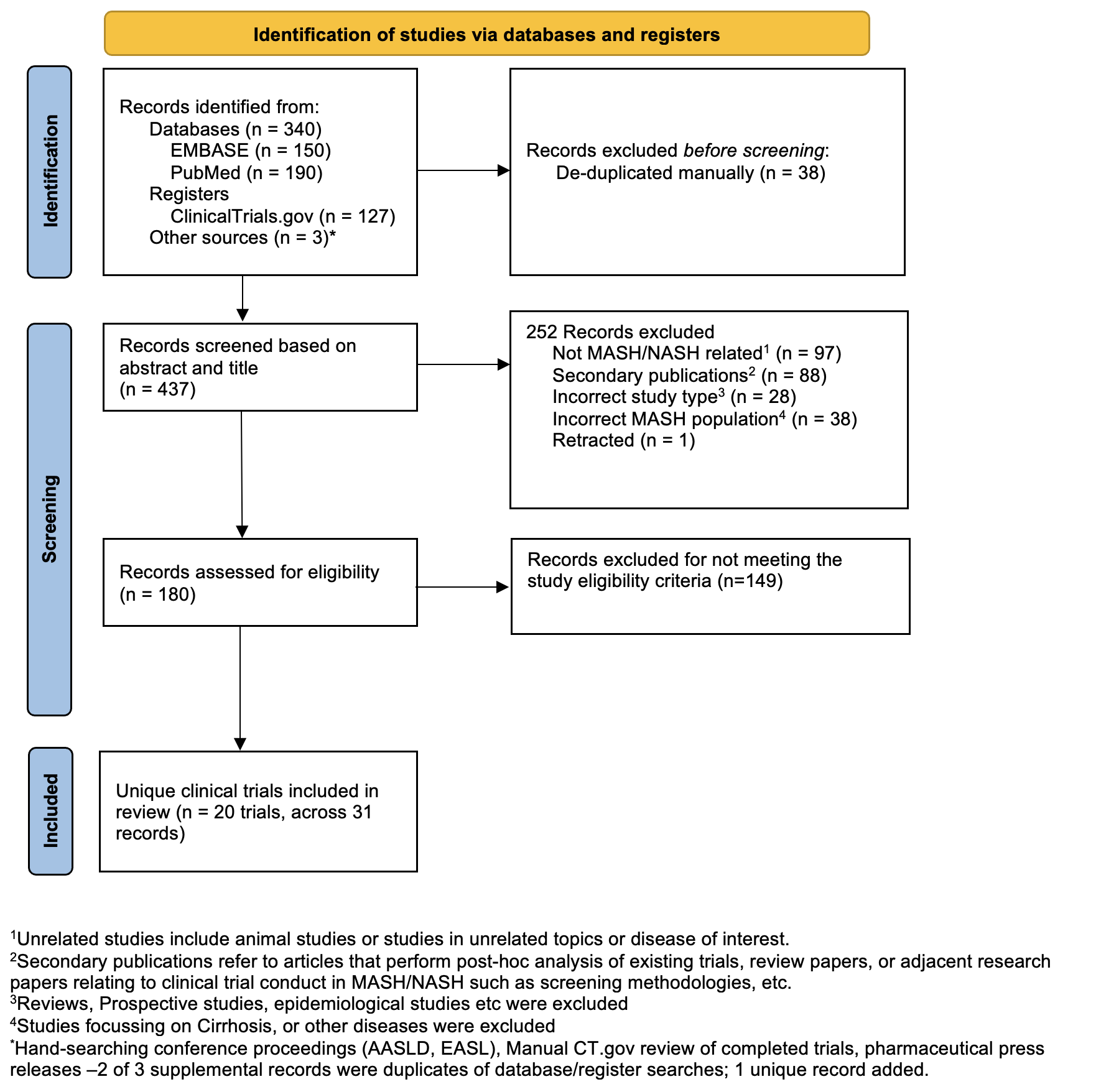

Supplementary Figure 5 **Quartile-based dose-response analysis of MRI-PDFF reduction and histological outcomes.** (A) NASH resolution and (B) fibrosis improvement across quartiles of relative MRI-PDFF change from baseline. Q1 denotes the greatest MRI-PDFF reduction and Q4 the smallest. Points show pooled rates and error bars show 95% confidence intervals from random-effects meta-analysis of logit-transformed proportions, back-transformed to percentages. Models accounted for within-trial clustering when multiple arms from the same study contributed to the same quartile. Quartile analyses were descriptive and used to complement the primary continuous meta-regression. There was no evidence for non-linearity by restricted cubic spline analyses.

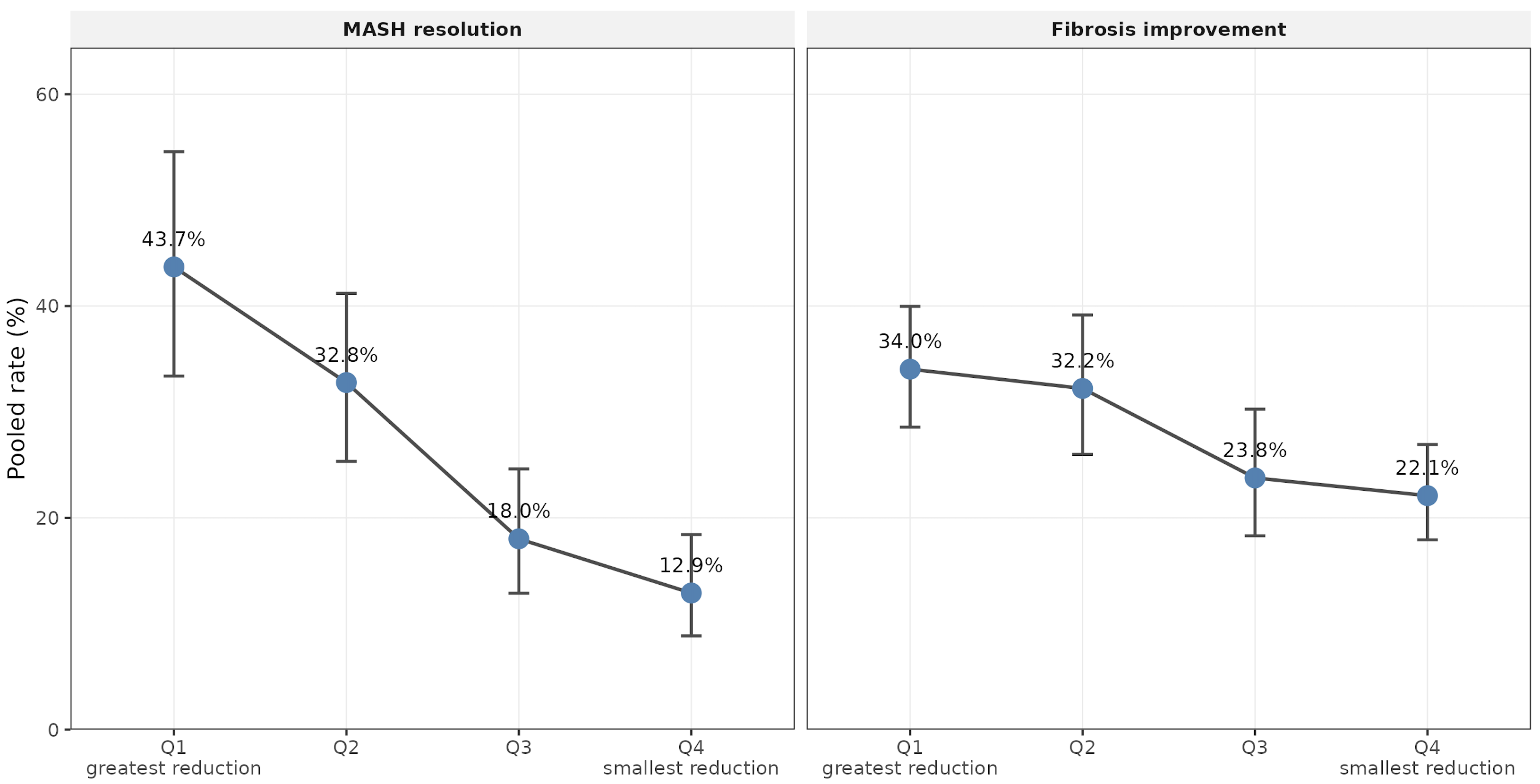

**Supplementary Figure 6 Leave one trial out sensitivity analyses**

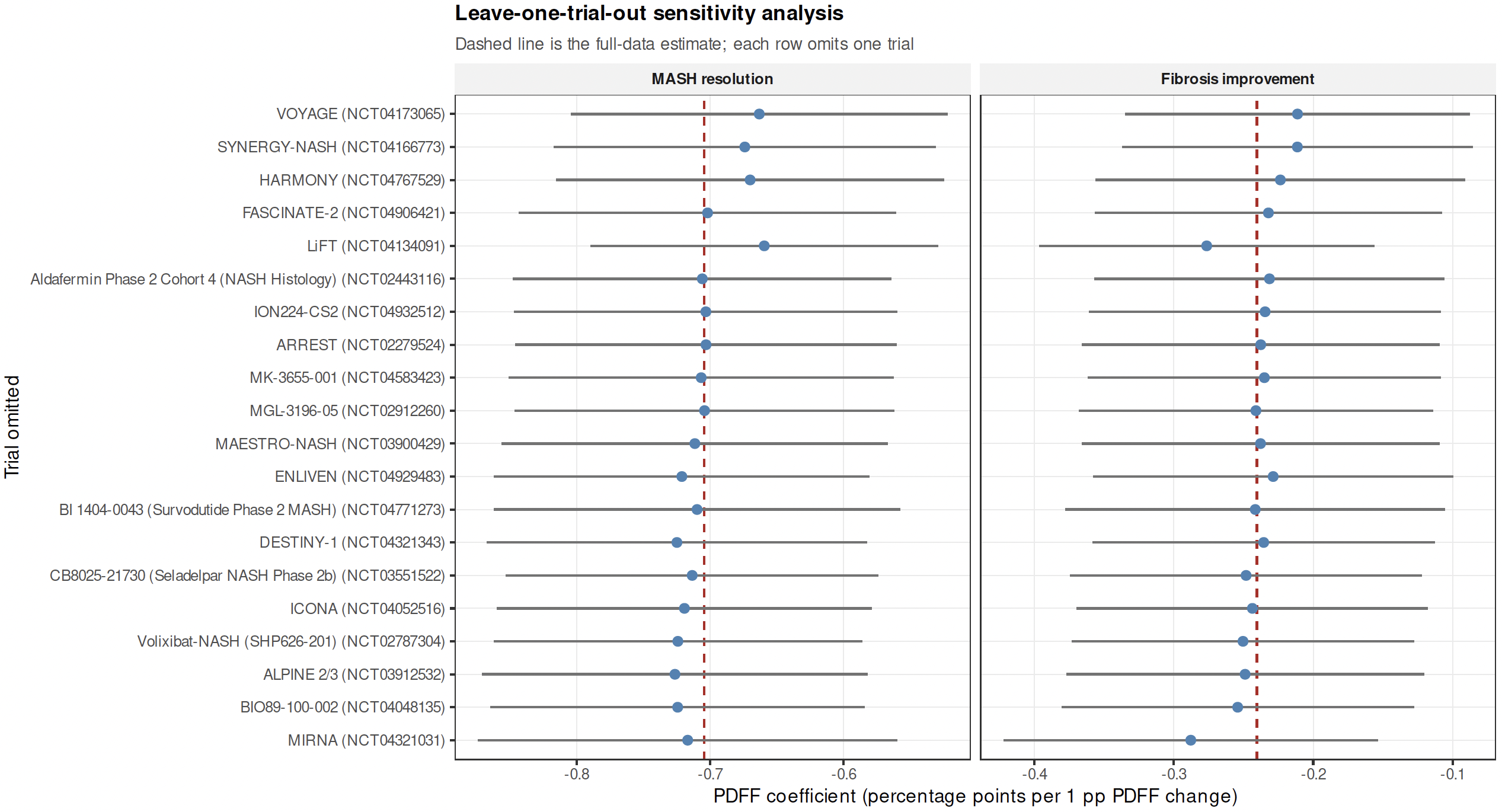

### Supplementary Notes

#### Notes on clinical studies used in the systematic meta-analysis

##### NCT03900429 - MAESTRO-NASH - THR-beta agonist

Notes:

- **PDFF data available at two timepoints per arm (wk 16, 52).** Histology only at Week 52 (single biopsy timepoint).
- No N for histology endpoints because the paper reports rates from a partial-credit CMH model (averaging two pathologists' reads), not simple event counts.
- ALT change is reported at Week 48 (not 52) and only for the subgroup with baseline ALT ≥30 U/L
- Absolute PDFF change (in percentage points) was not reported, only relative % change from baseline
- NAS total score at baseline wasn't reported as a continuous mean ± SD , only the proportion with NAS ≥5

##### NCT02443116 - Aldafermin Phase 2 Cohort 4 (NASH Histology) - FGF19 analog

Notes

- PDFF over time (4 timepoints per arm): Weeks 6, 12, 24, and 30 (off-drug).
- Histology at week 24 (paired biopsies: 50 aldafermin, 22 placebo).
- The week 24 PDFF data in Table 2 are reported as mean (SD), while CTgov and interim timepoints use LS mean (SE).
- Aldafermin development was subsequently discontinued after the ALPINE 2/3 and ALPINE 4 Phase 2b trials failed.

##### NCT04048135 - BIO89-100-002 - FGF21 analogue

Notes

- Part 2 is open-label with no placebo
- Baseline fibrosis stage distributions and NAS scores were not reported per arm in either the paper or CT.gov.
- Part 1 had no histology endpoints (no biopsies at end of treatment)
- The populations differ: Part 1 was mixed biopsy-confirmed + phenotypic NASH; Part 2 required biopsy-confirmed NASH with NAS≥4 and F2/F3

##### NCT04134091 – LiFT - Oral TU prodrug, Oral TU prodrug + VitE

Notes

- PDFF measured on 3 arms (Treatment A, Treatment B, Placebo) at 2 timepoints (Week 12, Week 36).
- Baseline PDFF per arm not reported; only change-from-baseline values available.
- Baseline ALT/AST per arm, only pooled all-group means reported in the press release (ALT ~51.5, AST ~31.9 U/L).
- BMI, weight, NAS score, fibrosis stage distribution per arm , not available from any public source.
- Diabetes prevalence per arm not reported.
- The fibrosis improvement endpoint (NASH CRN) paradoxically favored placebo (40%) over treatment arms (27%, 14%), which conflicts with the paired technique and FibroNest results that favored treatment.
- The study was male-only
- The study was not powered for histological endpoints
- Treatment B includes vitamin E (d-alpha tocopherol 476 mg/day).

##### NCT04583423 - MK-3655-001 - FGFR1c/KLB agonist

Notes

- PDFF data (Week 24 only): The relative change values are LS means from a constrained longitudinal data analysis (cLDA) model, not raw means.
- Histology data (Week 52) Only 73 of 183 randomized participants were included due to sponsor-initiated early termination.
- Missing data: Baseline ALT, AST, NAS scores, and absolute PDFF change are not reported by arm in any of the available sources.
- NAS reductions were back-calculated from the reported percentages and denominators (e.g., 33.3% of 18 = 6 events for 50 mg arm).

##### NCT04767529 – HARMONY - FGF21 analogue

Notes

- PDFF data available at two timepoints per arm, week 24 and 96.
- The placebo effect for PDFF absolute change is the LS mean difference from MMRM (from CTR analyses section). The relative % change vs placebo comes from the papers.
- Histology denominators are from the LBAS method (completers with paired biopsies) for all histology rates, which is the prespecified primary analysis. The MicroTally Mitt/mITT rates (missing=non-responder) are lower and available in the source documents (not recorded).

##### NCT04771273 - BI 1404-0043 (Survodutide Phase 2 MASH) - GCGR/GLP-1R dual agonist

Notes

- Planned maintenance treatment (ITT), consistent with the NEJM paper's main figures. This is more conservative than the "actual treatment" analysis (which was the protocol-specified primary) but more appropriate for meta-analysis since it preserves randomization.
- Week 28 PDFF data were collected but not yet published, the supplement mentions that these will be presented separately; only Week 48 is available.
- The 6.0 mg arm had the highest dropout (25/75), which dilutes the ITT results vs the actual-treatment analysis
- NASH resolution rates were read from supplement bar charts (Figure S6C) and the N events were back-calculated from rate x N
- ALT/AST SDs for change values were not individually extractable from the bar charts (only shown as error bars)

##### NCT04906421 - FASCINATE-2 - FASN inhibitor

Notes

- PDFF data extracted at two timepoints, 26 Wk and 52 Wk:
- No absolute PDFF change (in percentage points) is reported in the paper, only relative % change.
- Week 26 PDFF sample sizes aren't stated explicitly in the paper. The week 52 PDFF responder analysis used n=69 (denifanstat) and n=39 (placebo) among those with ≥8% baseline PDFF.
- NAS baseline scores are reported only categorically (4–5 vs 6–8), not as mean ± SD.

##### NCT04929483 – ENLIVEN - FGF21 analogue

Notes

- Only one timepoint (week 24). The paper mentions week 12 PDFF was a secondary endpoint, but individual arm-level week 12 PDFF data are not reported in the publication or supplement.
- Histologic outcomes used multiple imputation + stratified CMH. N events for histologic endpoints are not reported as raw counts in the imputed analysis (rates from multiple imputation don't correspond to integer counts). The completer analysis in Table S5/S6 does give observed counts (e.g., 4/54 placebo, 14/53 for 30 mg QW for fibrosis improvement).

##### NCT02279524 – ARREST - SCD1 inhibitor

Notes

- ARREST used MRS (magnetic resonance spectroscopy) measuring the triglyceride-to-water ratio, not MRI-PDFF.
- The ALT/AST longitudinal curves (weeks 24, 40, 52) are shown in Figure 2 of the paper but exact values at intermediate timepoints are not tabulated, so only week 52 LS means could be extracted.
- Relative PDFF change , not reported; only absolute change in MRS liver fat % was provided
- Fibrosis stages F0 and F1 by arm , not broken out per arm in the paper (only F2 and F3 percentages reported per arm)
- Baseline PDFF samplesize uses FAS MRI subset, which is smaller than randomized N due to missing paired scans
- Responder n events for ≥30% relative reduction calculated from the reported percentages x denominators (e.g., 30.1% × 83 ≈ 25 for 600 mg arm).

##### NCT02912260 - MGL-3196-05 - THR-beta agonist

Notes

- Two arms (Placebo and pooled Resmetirom) at two timepoints (week 12 and week 36), values from the Lancet paper's Tables 2,5 and Supplementary Table S1. Week 36 matched with biopsy.
- **Dose heterogeneity,** this was an adaptive dosing study, 47% of patients were reduced to 60mg at week 4. The "all doses" arm is the ITT-equivalent wich was recorded. The post-hoc 60mg vs 80mg split in Appendix Table S2 shows the 80mg group had much stronger PDFF reduction (–50.2% relative) and NASH resolution (39.4%) than 60mg (–39.4% and 19.4%). The dose was labeled 80mg since that was the assigned dose.
- **NASH resolution denominator**: The prespecified analysis excluded patients with ≥9.5% weight loss (n=31 placebo, n=73 resmetirom). The "including weight loss >9.5%" result was also reported (14.7% vs 24.7%, non-significant).
- **No per-arm baseline split** is available for the dose subgroups, only the pooled resmetirom arm baseline was reported in Table 1.

##### NCT03551522 - CB8025-21730 (Seladelpar NASH Phase 2b) - PPARδ agonist

**Notes**

- No dedicated publication was found for this trial. The study was terminated due to unexpected histological findings (later deemed pre-existing).
- The Week 12 PDFF data uses LS means (ANCOVA), while Week 52 PDFF uses arithmetic mean/SD
- Baseline PDFF, ALT, AST, BMI, weight, NAS, and fibrosis distribution by arm are **not reported** in the ClinicalTrials.gov results (only overall summaries from the press release: mean PDFF ~21%, ALT ~62, AST ~46, NAS ~5.2, ~83% F2-F3)
- Notably, the placebo arm showed the largest PDFF reduction at Week 12 (LS mean −20.78%) , greater than all seladelpar doses.
- **Missing per-arm baseline data**: ClinicalTrials.gov does not report baseline PDFF, ALT, AST, BMI, weight, NAS, fibrosis stage distribution, or diabetes prevalence broken out per arm, only pooled overall values from the press release.
- **Week 52 PDFF statistic change**: The Week 12 primary endpoint used LS means (ANCOVA), but Week 52 PDFF was reported as arithmetic mean ± SD.
- **Early termination**: The study was terminated due to "unexpected histological findings" (later exonerated), so the Week 52/ET data includes some subjects who had their biopsy at the early termination.

##### NCT04932512 - ION224-CS2 - DGAT2i (ASO)

Notes

- Absolute PDFF change was not reported numerically, only relative change LSM is available.
- 60 mg arm was not expanded and has limited data (n=18 PPS); histology endpoint rates were not formally reported in the main figures.
- Baseline characteristics are from the ITT population (n=46/23/45/46), while efficacy data are from PPS (n=32/18/39/34).

##### NCT04166773 - SYNERGY-NASH - GIP/GLP-1 RA

Notes

- SYNERGY-NASH only reports PDFF at a single timepoint (Week 52). No interim PDFF data (e.g., Week 12, 24, 26) were published in the paper, supplement, or CTgov results. The interim futility analysis used Week 26 PDFF data internally, but those values were never disclosed.
- The paper (Figure 1) reports NASH resolution rates of 44/56/62/10% while CTgov reports 51.84/63.13/73.92/12.62% , the CTgov values are from the per-protocol-like efficacy analysis set (n=39/41/40/35 with evaluable biopsies), while the paper values incorporate MI across the full ITT population (N=47/47/48/48). For the meta-analyses, the paper is rates are used.
- Similarly for fibrosis improvement: paper 55/51/51/30% vs CTgov 59.21/53.35/54.30/32.51%.

##### NCT04321031 – MIRNA - DGAT2i, DGAT2i + ACCi

Notes

- Only week 48 PDFF is numerically reported in the CT.gov results and the paper's supplementary Table S4. The paper's Figure 3A shows week 6 and 24 graphically but doesn't provide exact numbers in text for all arms at those timepoints. The study completed with only 73% enrollment due to COVID-19
- PDFF was only measured in a North American MRI-PDFF substudy (~113 enrolled, 78 completed), so sample sizes per arm at week 48 are very small (n=8–18). The ANCOVA LS mean percent change values and SE are from CT.gov results. The vs-placebo differences are the LS mean differences from the ANCOVA model reported on CT.gov.
- The paper also provides Bayesian model estimates for PDFF (e.g., placebo: –10.79%, E300: –55.53%) but ANCOVA pairwise results are recorded since those are more standard for meta-analysis.
- PDFF was also measured at weeks 6 and 24, but exact numeric values per arm are only shown graphically in the paper (Figure 3A), not in tabular form. If you need those intermediate timepoints, they would need to be extracted from the published figure or the appendix data tables.
- All histology is at week 48 only (biopsy at screening and week 48). Non-responder imputation was used for missing biopsies.
- All treatment differences and CIs are 90% CIs (not 95%), per the study's pre-specified estimation approach.
- Missing week 48 biopsies were imputed as non-responders in primary analysis
- No absolute PDFF change data reported (only relative % change)
- No interim timepoint PDFF values available.
- Baseline PDFF only for the substudy subset, not the full population
- High placebo response for fibrosis improvement (35.3%) may have confounded results

##### NCT04321343 - DESTINY-1 - deuterated TZD (R-pio)

Notes

- For PDFF data, the primary analysis used ANCOVA LS means with multiple imputation (MAR) for the ITT set (n=117). The vs. placebo effect for relative PDFF change is the LS mean difference from that ANCOVA model.
- Histology data: Fibrosis improvement and NASH resolution were analyzed in biopsy completers only (n=92 total: 21, 22, 26, 23 per arm), not the full ITT set.
- No individual fibrosis stages reported, only as F1 vs. F2/F3 combined (stratification groups).

##### NCT04173065 – VOYAGE - THR-beta agonist

Notes

- 2 PDFF times points available: Median PDFF on Week 12 (from the AASLD oral), and Week 52 PDFF from press release is LS mean (ANCOVA). The topline press release at Week 12 reported median range 38%–55% without arm-level LS means.
- No SD/SE/CI reported for PDFF changes in any source, only point estimates and p-values vs placebo.
- N events for histology were back-calculated from rates x denominators (from the press release tables).

### Ethics Statements

#### UK Biobank

Ethical approval for the UK Biobank was previously obtained from the North West Centre for Research Ethics Committee (11/NW/0382). The work described herein was approved by the UK Biobank under application number 26041. Informed consent was obtained for all study participants.

#### GHS-RGC DiscovEHR Collaboration

The MyCode Community Initiative parent study was approved by the Geisinger IRB (Study #2006-0258). The MyCode Governing Board reviews and approves all uses of MyCode samples and data. Additionally, the Geisinger IRB reviewed this study and determined the study did not involve human subjects as defined in 45 CFR 46.102(f); and therefore, was not subject to additional oversight by the IRB (Study #2017-158).

#### Mayo Clinic Project Generation

The study was approved by the Mayo Clinic Institutional Review Board under protocol number 19-007763 and is compliant with the ethical guidelines of the Helsinki and Istanbul Declarations.

#### Colorado Center for Personalized Medicine

Biospecimens and associated data used in this study were obtained from the biobank at the Colorado Center for Personalized Medicine (CCPM) at the University of Colorado Anschutz Medical Campus (CU AMC). All samples and data were collected under Institutional Review Board (IRB) approved protocol (#15-0461) with appropriate informed consent from participants. Research using these materials was conducted in accordance with the ethical guidelines and regulations governing human subjects research, upholding the principles of respect for persons, beneficence, and justice.

#### UCLA-RGC ATLAS Collaboration

All individuals provided written informed consent to participate in the study. Patient Recruitment and Sample Collection for Precision Health Activities at UCLA is an approved study by the UCLA Institutional Review Board (UCLA IRB). IRB#17-001013.

#### Mount Sinai Million Health Discoveries Program

The Icahn School of Medicine at Mount Sinai's IRB, Program for the Protection of Human Subjects (PPHS), approved the BioMe Biobank and Mount Sinai Million Health Discoveries Program (PPHS IRB #11-01139 and #21-01743).

#### Penn Medicine Biobank

The PMBB is approved under IRB protocol #813913 and supported by Perelman School of Medicine at University of Pennsylvania, a gift from the Smilow family, and the National Center for Advancing Translational Sciences of the National Institutes of Health under CTSA award number UL1TR001878.

#### Malmö Diet and Cancer Study

IRB approval for the Malmö Diet and Cancer Study was obtained from the Ethics Committee of Lund University (LU 51-90) and the Regional Board of Ethics in Lund (Dnr 2016/479).

#### Indiana University School of Medicine (Indiana-CLDB)

Indiana Biobank protocol has been reviewed by IU's IRB and approved under the protocol number 1105005445. All participants have provided written informed consent.

#### NASH CRN

Individuals included in this study participated in the NASH CRN DR1, DB2, and DB3, PIVENS, FLINT, TONIC, and CyNCh trials at multiple adult and pediatric clinical centers; the data coordinating center was located at the Johns Hopkins University. A data safety and monitoring board (DSMB) appointed and managed by the NIDDK provided oversight for all studies of the NASH CRN. Each protocol was approved at every participating center and all participants provided written informed consent.

### Acknowledgements

#### Regeneron Genetics Center

RGC Management & Leadership Team
Aris Baras, Gonçalo Abecasis, Adolfo Ferrando, Giovanni Coppola, Andrew Deubler, Luca A Lotta, John D Overton, Jeffrey G Reid, Alan Shuldiner, Katherine Siminovitch, Jason Portnoy, Marcus B Jones, Lyndon Mitnaul, Alison Fenney, Jonathan Marchini, Manuel Allen Revez Ferreira, Maya Ghoussaini, Mona Nafde, William Salerno, Cristen Willer, Lourdes Crane, Niek Verweij, Eric Jorgenson, and Joseph Pickrell.

*Sequencing & Lab Operations*
John D Overton, Christina Beechert, Erin Fuller, Laura M Cremona, Eugene Kalyuskin, Hang Du, Caitlin Forsythe, Zhenhua Gu, Kristy Guevara, Michael Lattari, Alexander Lopez, Kia Manoochehri, Prathyusha Challa, Manasi Pradhan, Raymond Reynoso, Ricardo Schiavo, Maria Sotiropoulos Padilla, Chenggu Wang, Sarah E Wolf, Hang Du, Kristy Guevara.

*Genome Informatics & Data Engineering*
Jeffrey G Reid, Mona Nafde, Manan Goyal, George Mitra, Sanjay Sreeram, Rouel Lanche, Vrushali Mahajan, Sai Lakshmi Vasireddy, Gisu Eom, Krishna Pawan Punuru, Sujit Gokhale, Shehroze Aamer, Pooja Mule, Mudasar Sarwar, Muhammad Aqeel, Xiaodong Bai, Lance Zhang, Sean O'Keeffe, Razvan Panea, Evan Edelstein, Devika Torvi, Ayesha Rasool, William Salerno, Evan K Maxwell, Boris Boutkov, Alexander Gorovits, Ju Guan, Alicia Hawes, Olga Krasheninina, Samantha Zarate, Adam J Mansfield, Lukas Habegger, Stephen Tahan, Naveen Karumuri.

*Analytical Genetics and Data Science*
Gonçalo Abecasis, Manuel Allen Revez Ferreira, Joshua Backman, Kathryn Burch, Adrian Campos, Liron Ganel, Sheila Gaynor, Benjamin Geraghty, Arkopravo Ghosh, Christopher Gillies, Lauren Gurski, Eric Jorgenson, Tyler Joseph, Michael Kessler, Jack Kosmicki, Adam Locke, Priyanka Nakka, Jonathan Marchini, Karl Landheer, Olivier Delaneau, Maya Ghoussaini, Anthony Marcketta, Joelle Mbatchou, Jonathan Ross, Carlo Sidore, Eli Stahl, Timothy Thornton, Rujin Wang, Kuan-Han Wu, Bin Ye, Blair Zhang, Andrey Ziyatdinov, Yuxin Zou, Jingning Zhang, Kyoko Watanabe, Mira Tang, Frank Wendt, Suganthi Balasubramanian, Suying Bao, Kathie Sun, Chuanyi Zhang, Sean Yu, Aaron Zhang, David Corrigan, Dhruv Shidhaye, Chen Wang, Keyrun Adhikari, Alexander Lachmann, Anna Alkelai, Mark Weiner, Julian Stamp.

*Therapeutic Area Genetics*
Adolfo Ferrando, Giovanni Coppola, Luca A. Lotta, Alan Shuldiner, Katherine Siminovitch, Brian Hobbs, Jon Silver, William Palmer, Rita Guerreiro, Amit Joshi, Antoine Baldassari, Cristen Willer, Sarah Graham, Ernst Mayerhofer, Erola Pairo Castineira, Mary Haas, Niek Verweij, George Hindy, Jonas Bovijn, Tanima De, Luanluan Sun, Olukayode Sosina, Arthur Gilly, Peter Dornbos, Moeen Riaz, Manav Kapoor, Gannie Tzoneva, Veera Rajagopal, Sahar Gelfman, Vijay Kumar, Jacqueline Otto, Jose Bras, Silvia Alvarez, Jessie Brown, Hossein Khiabanian, Joana Revez, Kimberly Skead, Jae Soon Sul, Lei Chen, Sam Choi, Amy Damask, Nan Lin, Charles Paulding, Sameer Malhotra, Joseph Herman, Jacob McPadden, David Blair, Joshua Motelow, Julie Horowitz.

*Research Program Management & Strategic Initiatives*
Marcus B Jones, Michelle G LeBlanc, Nadia Rana, Jennifer Rico-Varela, Jaimee Hernandez, Larizbeth Romero, Ashley Paynter.

*Senior Partnerships & Business Operations*

Randi Schwartz, Lourdes Crane, Alison Fenney, Jody Hankins, Anna Han, Samuel Hart, Ryan Smith, Sarah Murphy.

*Business Operations & Administrative Coordinators*

Ann Perez-Beals, Gina Solari, Johannie Rivera-Picart, Michelle Pagan, Sunilbe Siceron.

Affiliations:

Regeneron Genetics Center, Tarrytown, NY, USA.

For RGC contact info, please use: 

#### GHS-RGC DiscovEHR Collaboration

- Adam Buchanan. Geisinger Health System, Danville, PA, USA
  -
- David J. Carey. Geisinger Health System, Danville, PA, USA
  -
- Christa L. Martin. Geisinger Health System, Danville, PA, USA
  -
- Michelle Meyer. Geisinger Health System, Danville, PA, USA
  -
- Kyle Retterer. Geisinger Health System, Danville, PA, USA
  -
- David Rolston. Geisinger Health System, Danville, PA, USA
  -

#### Colorado Center for Personalized Medicine

Heather D. Anderson, Christina L. Aquilante, Kelsey Arbogast, Ian M. Brooks, Elizabeth E. Burke, Emily M. Casteel, Joanne B. Cole, Curtis R. Coughlin II, Jacob Crawford, Kristy Crooks, Erin Culver, Matthew J. Fisher, Teresa C. Frye, Hunter George,

Chris R. Gignoux, Elizabeth K. Gilliland, Casey S. Greene, Emily Hearst, Audrey E. Hendricks, Randi K. Johnson, Shelby Jones, Dave Kao, Gabrielle A. Knortz, Danielle Koffenberger, Santhanagopalan Krishnamoorthy, Lisa Ku, Elizabeth L. Kudron, Rashawnda Lacy, Ethan M. Lange, Joe A. Lesny, Meng Lin, James L. Martin.

For CCPM contact info, please use:

#### Penn Medicine Biobank

*PMBB Leadership Team*

Daniel J. Rader, M.D., Marylyn D. Ritchie, Ph.D.

Contribution: All authors contributed to securing funding, study design and oversight. All authors reviewed the final version of the manuscript.

*Patient Recruitment and Regulatory Oversight*

JoEllen Weaver, Nawar Naseer, Ph.D., M.P.H., Giorgio Sirugo, M.D., P.h.D., Afiya Poindexter, Yi-An Ko, Ph.D., Kyle P. Nerz, Jenna Dever, Aidan Harvey, Sydney Linn

Contributions: JW manages patient recruitment and regulatory oversight of study. NN manages participant engagement, assists with regulatory oversight, and researcher access. GS assists with researcher access. AP, YK, KPN, JD, AH, and SH perform recruitment and enrollment of study participants.

*Lab Operations*

JoEllen Weaver, Meghan Livingstone, Fred Vadivieso, Stephanie DerOhannessian, Teo Tran, Julia Stephanowski, Salma Santos, Ned Haubein, P.h.D., Joseph Dunn

Contribution: JW, ML, FV, SD conduct oversight of lab operations. ML, FV, AK, SD, TT, JS, SS perform sample processing. NH, JD are responsible for sample tracking and the laboratory information management system.

*Clinical Informatics*

Anurag Verma, Ph.D., Colleen Morse Kripke, M.S. DPT, MSA, Marjorie Risman, M.S., Renae Judy, B.S., Colin Wollack, M.S.

Contribution: All authors contributed to the development and validation of clinical phenotypes used to identify study subjects and (when applicable) controls.

*Genome Informatics*

Anurag Verma Ph.D., Shefali S. Verma, Ph.D., Scott Damrauer, M.D., Yuki Bradford, M.S., Scott Dudek, M.S., Theodore Drivas, M.D., Ph.D.,

Contribution: AV, SSV, and SD are responsible for the analysis, design, and infrastructure needed to quality control genotype and exome data. YB performs the analysis. TD and AV provide variant and gene annotations and their functional interpretation of variants.

For PMBB contact info, please use: 

#### Mayo Clinic Project Generation

**Mayo Clinic-RGC Project Generation (PG)**

*PG Leadership Team*

Cerhan, James R., M.D., Couch, Fergus J., Ph.D., Olson, Janet E., Ph.D.

*Statistical Genetics and Bioinformatics*

Larson, Nicholas B., Ph.D., M.S., Fredericksen, Zachary S.

*Laboratory Operations*

Cicek, Mine, Ph.D.

*Registry Principal Investigators*

(Alphabetical listing)

1. Alcohol Use Disorder (AUD): Biernacka, Joanna M., Ph.D., Karpyak, Victor M., M.D. Ph.D.
2. Alzheimer's Disease Research Center (ADRC): Vemuri, Prashanthi, Ph.D., Ramanan, Vijay K., M.D., Ph.D., Ross, Owen A., Ph.D.
3. Bipolar disorder registry: Biernacka, Joanna M., Ph.D., Frye, Mark A., M.D.
4. Brain: Eckel Passow, Jeanette E., Ph.D., Jenkins, Robert R., M.D., Ph.D., Lachance, Daniel H., M.D., Drucker, Kristen L, Ph.D., Decker, Paul A., M.S., Kosel, Matthew L.
5. Breast - Mayo Florida: McLaughlin, Sarah A., M.D.
6. Breast - Mayo Rochester: Olson, Janet E., Ph.D.; Couch, Fergus J., Ph.D., Ruddy, Kathryn J., M.D., Boddicker, Nicholas J., Ph.D., Chen, Wenan, Ph.D.
7. Cardiovascular Disease Specimen Repository: Bielinski, Suzette J., Ph.D., M.Ed.
8. Chronic Kidney Disease & Kidney Stone Disease: Lieske, John C., M.D.
9. Chronic Pain: Hooten, W. Michael, M.D.
10. Colorectal: Boardman, Lisa A., M.D.
11. COVID-19 Biobank: Kennedy, Richard B., Ph.D., Cerhan, James R., M.D., Ph.D., Badley, Andrew D., M.D.
12. Endometrial: Dowdy, Sean C., M.D., Bakkum-Gamez, Jamie N., M.D., Harrington, Shariska, M.D., Glaser, Gretchen E., M.D.
13. Lung: Yang, Ping, M.D., Ph.D.
14. Lymphoma: Cerhan, James R., M.D., Ph.D.
15. Mayo Clinic Biobank: Olson, Janet E., Ph.D.
16. Mayo Clinic Study of Aging (MCSA): Vemuri, Prashanthi, Ph.D., Ramanan, Vijay K., M.D., Ph.D.; Ross, Owen A., Ph.D.
17. Mayo Mammography Health Study: Vachon, Celine M., Ph.D., Stacey Winham Ph.D.
18. Multiple Myeloma (MM)/Smouldering MM (SMM): Dispenzieri, Angela, M.D., Vachon, Celine M., Ph.D.
19. Neuroendocrine pancreatic tumor registry: Antwi, Samuel O., Ph.D., Ann L. Oberg, Ph.D., Kari G. Rabe, MS.
20. Mayo Clinic Biospecimen Resource for Ovarian Cancer Research: Kaufmann, Scott H., M.D., Ph.D., Goode, Ellen L., Ph.D., William A. Cliby, M.D., Jamie Bakkum-Gamez, M.D., Sun-Hee Lee, Ph.D., Stacey Winham Ph.D.
21. Biospecimen Resource for Pancreas Research: Antwi, Samuel O., Ph.D.; Ann L. Oberg, Ph.D.; Kari G. Rabe, MS.
22. Parkinson’s Disease: Ahlskog, J. Eric, M.D., Ph.D.; Bower, James H., M.D.
23. Polycystic kidney disease: Harris, Peter C., Ph.D.
24. Polyps: Boardman, Lisa A., M.D.
25. Prevalence of Asymptomatic Ventricular Dysfunction: Pereira, Naveen L., M.D.
26. PRISM Mammography study: Couch, Fergus J., Ph.D.; Vachon, Celine M., Ph.D.; Olson, Janet E., Ph.D.
27. Prostate: Cicek, Mine, Ph.D.
28. Prostate Family: Cicek, Mine, Ph.D.
29. 2Radiation Oncology Registry: Laack, Nadia N., M.D.; Ma, Daniel J., M.D.; Mutter, Robert W., M.D.
30. Renal: Antwi, Samuel O., Ph.D.; Eckel Passow, Jeanette E., Ph.D.
31. Vascular Diseases Biorepository: Pereira, Naveen L., M.D.

*Management*

Harrington, Jonathan J.

For Mayo-Clinic contact info, please use both e-mail addresses:

1. Couch, Fergus J., Ph.D.
2. Cerhan, James R., M.D., Ph.D.

**MAYO-RGC Project Generation – Financial Support**

*Registry Principal Investigators*

(Alphabetical listing)

- General Support: Mayo Clinic Center for Individualized Medicine and Mayo Clinic Comprehensive Cancer Center (P30 CA15083), and Mayo Clinic Foundation
- Alcohol Use Disorder (AUD): Biernacka, Joanna M., Ph.D.; Karpyak, Victor M., M.D., Ph.D.
  - NIAAA (P20 AA017830) and the Samuel C Johnson Genomics of Addiction Program at Mayo Clinic.
- Alzheimer's Disease Research Center (ADRC): Ramanan, Vijay K., M.D., Ph.D.; Ross, Owen A., Ph.D.; Vemuri, Prashanthi, Ph.D.
  - Alzheimer's Disease Research Center (ADRC), P30 AG062677
- Bipolar disorder registry: Biernacka, Joanna M., Ph.D.; Frye, Mark A., M.D.
  - J. Willard and Alice S. Marriott Foundation.
- Brain: Jenkins, Robert B., M.D., Ph.D.; Eckel Passow, Jeanette, E., Ph.D.
  - NIH R01 CA230712, U19CA264362
- Breast - Mayo Florida: McLaughlin, Sarah A., M.D.
  - NIH R21 CA191270
  - Bankhead-Coley Cancer Research Program (2BN01)
- Breast _Mayo Rochester: Olson, Janet E., Ph.D.; Couch, Fergus J., Ph.D.
  - NIH/NCI R35 CA253187 and P50 CA116201
- Cardiovascular Disease Specimen Repository: Bielinski, Suzette J., Ph.D., M.Ed.
- Chronic Kidney Disease & Kidney Stone Disease: Lieske, John C., M.D.

NIH R01 DK133171

- Chronic Pain: Hooten, W. Michael, M.D.
- Colorectal: Boardman, Lisa A., M.D.
- COVID-19 Biobank: Kennedy, Richard B., Ph.D.; Cerhan, James R., M.D., Ph.D.; Badley, Andrew D., M.D.
- Endometrial: Dowdy, Sean C., M.D.; Bakkum-Gamez, Jamie N., M.D.; Glaser, Gretchen E., M.D.
- Lung: Yang, Ping, M.D., Ph.D.
- Lymphoma: Cerhan, James R., M.D., Ph.D.
  - NIH/NCI P50 CA97274, R01 CA92153, Predolin Foundation
- Mayo Clinic Biobank: Olson, Janet E., Ph.D.
  - Mayo Clinic Center for Individualized Medicine.
- Mayo Clinic Study of Aging (MCSA): Vemuri, Prashanthi, Ph.D.; Ramanan, Vijay K., M.D., Ph.D.; Ross, Owen A., Ph.D.
  - NIH U01 AG006786; R01 AG56366; GHR foundation
- Mayo Mammography Health Study: Vachon, Celine M., Ph.D; Winham, Stacey Ph.D.
  - NIH/NCI R01 CA128931; R01 CA97396; Mayo Clinic Cancer Center
- Multiple Myeloma (MM)/Smouldering MM (SMM): Dispenzieri, Angela, M.D.; Vachon, Celine M., Ph.D.;
  - NIH/NCI R01 CA271014
- Neuroendocrine pancreatic tumor registry: Antwi, Samuel O., Ph.D.; Ann L. Oberg, Ph.D.
  - NIH/NCI P50CA102701, and Mayo Clinic Foundation
- Ovarian: Kaufmann, Scott H., M.D., Ph.D.; Goode, Ellen L., Ph.D.
  - NIH/NCI P50 CA136393
- Biospecimen Resource for Pancreas Research: Antwi, Samuel O., Ph.D.; Ann L. Oberg, Ph.D.
  - NIH/NCI P50CA102701, and Mayo Clinic Foundation
- Parkinson’s Disease: Ahlskog, J. Eric, M.D., Ph.D.; Bower, James H., M.D.
- Polycystic kidney disease: Harris, Peter C., Ph.D.
  - NIH R01 DK058816; DK059597
- Polyps: Boardman, Lisa A., M.D.
- Prevalence of Asymptomatic Ventricular Dysfunction: Pereira, Naveen L., M.D.
- PRISM Mammography study: Couch, Fergus J., Ph.D.; Vachon, Celine M., Ph.D.; Olson, Janet E., Ph.D.
- Prostate: Mine Cicek, Ph.D.
- Prostate Family: Cicek, Mine, Ph.D.
- Radiation Oncology Registry: Laack, Nadia N., M.D.; Ma, Daniel J., M.D.; Mutter, Robert W., M.D.;
- Renal: Antwi, Samuel O., Ph.D.; Eckel Passow, Jeanette E., Ph.D.; Thompson, R. Houston, M.D.; Sharma, Vidit, M.D.
  - NIH/NCI R01CA134466 & Mayo Clinic Foundation
- Vascular Diseases Biorepository: Kullo, Iftikhar J., M.D.

#### Malmö Diet and Cancer Study

Olle Melander.

Department of Clinical Science Malmö, Lund University, Malmö, Sweden

Department of Internal Medicine, Skane University Hospital, Malmö, Sweden

#### NASH CRN

The Nonalcoholic Steatohepatitis Clinical Research Network (NASH CRN) is an NIDDK funded consortium and it conducted prospective studies and they collectively constitute the NASH CRN cohort included in this paper. All studies were approved by local IRB and all participants signed a written informed consent/ Briefly, patients with biopsy-confirmed MASLD were prospectively recruited at multiple medical centers across the United States between 2004 and 2020. The diagnosis of MASLD was based on >5% of hepatocytes containing macrovesicular steatosis and exclusion of significant alcohol consumption (>20g/d for women, >30g/d for men) within 2 years of the initial biopsy. All liver biopsies were reviewed in a blinded fashion by the NASH CRN Pathology Committee and scored according to the NASH CRN Scoring System. This collaboration follows by the NIH Genomic Data Sharing policies and procedures. As part of this collaboration, the RGC conducted whole exome sequencing at no-cost to the NASH CRN and returned results to the NASH CRN.

##### Financial support

This study was approved by the Nonalcoholic Steatohepatitis Clinical Research Network as an Ancillary Study (NASH CRN AS 116). The NASH CRN is supported by the National Institute of Diabetes and Digestive and Kidney Diseases (NIDDK) (U01DK061713, U01DK061718, U01DK061728, U01DK061731, U01DK061732, U01DK061734, U01DK061737, U01DK061738, U01DK061730, and U24DK061730). No funding was received from the NASH CRN for conducting this ancillary study. The sequencing was done in-kind by the Regeneron Genetics Center through a collaborative agreement.

The PIVENS trial was conducted by the NASH CRN and supported in part by Takeda Pharmaceuticals North America through a Cooperative Research and Development Agreement with the NIDDK. The vitamin E and matching placebo for the PIVENS trial were provided by Pharmavite through a Clinical Trial Agreement with the NIH. The FLINT trial was conducted by the NASH CRN and supported in part by a Collaborative Research and Development Agreement (CRADA) between NIDDK and Intercept Pharmaceuticals. The TONIC trial was conducted by the NASH CRN and supported in part by the Intramural Research Program of the National Cancer Institute and the Eunice Kennedy Shriver National Institute of Child Health and Human Development. The vitamin E and matching placebo were provided by Pharmavite through a Clinical Trial Agreement with the NIH. The CyNCh trial was conducted by the NASH CRN and supported in part by the Intramural Research Program of the National Cancer Institute and by a Collaborative Research and Development Agreement (CRADA) between NIDDK and Raptor Pharmaceuticals.

Ethics Statement

Individuals included in this study participated in the NASH CRN DR1, DB2, and DB3, PIVENS, FLINT, TONIC, CyNCn trial at multiple adult and pediatric clinical centers and the data coordinating center was located at the Johns Hopkins University. A data safety and monitoring board (DSMB) appointed and managed by the NIDDK provided oversight for all studies of the NASH CRN. Each protocol was approved at every participating center and all participants have provided a written informed consent.

Data Availability:

All study phenotype data and Biosamples are stored at the NIDDK Central Repository, (<https://repository.niddk.nih.gov/home/> ). Interested investigators should go to the website and complete the request forms and agreements to use the study data and/or request biospecimens for each NASH CRN study used in this manuscript. Sequencing data from this study will be deposited into public domain per the NIH Genomic Data Sharing Policy.

##### Acknowledgments

The authors thank the National Institute of Diabetes and Digestive and Kidney Diseases (NIDDK) for its support of the NASH CRN and this research. However, the content is solely the responsibility of the authors and does not necessarily represent the official views of the National Institutes of Health. The authors thank the Nonalcoholic Steatohepatitis Clinical Research Network (NASH CRN) investigators and the Ancillary Studies Committee for providing clinical samples and relevant data from the Nonalcoholic Fatty Liver Disease (NAFLD) Databases 1 and 2 (Adult and Pediatric), PIVENS, FLINT, TONIC, CyNCh and STOP-NAFLD trials. The authors also thank the participants involved in the NASH CRN studies. The biospecimens from the NASH CRN reported here were supplied by the NIDDK Central Repository. This manuscript was not prepared in collaboration with the NIDDK Central Repository and does not necessarily reflect the opinions or views of the NIDDK Central Repository.

#### Indiana University School of Medicine (Indiana-CLDB)

1) Participants from the Indiana Biobank cohort included in this analysis were recruited during January 1990 and June 2017.
2) **Ethics Statement:** Indiana Biobank protocol has been reviewed by IU's IRB and approved under the protocol number 1105005445. All participants have provided a written informed consent.
3) **Data Availability:** Data can be made available for those requesting via Indiana Biobank's data request process. Application is reviewed by three members of the Indiana Biobank’s Scientific Review Committee and if approved, made available on a secure-enclave for analysis.
4) **Acknowledgment:** This project was made possible, in part, with support from the Indiana Biobank and the Indiana Clinical and Translational Sciences Institute funded, in part by, grant number UM1TR004402 and the Lilly Endowment.”
5) **Financial disclosure:** Drs. Chalasani and Schwantes-An declare no conflict of interest for this paper.

Naga P. Chalasani.

Linus; Tae-Hwi L. Schwantes-An.

Andrew J. Saykin.

All authors are affiliated to Indiana University School of Medicine, Indianapolis, IN, USA.

#### Mount Sinai Million Health Discoveries Program

Alexander W. Charney, MD

Affiliation: Icahn School of Medicine at Mount Sinai

#### UCLA-RGC ATLAS Collaboration

| **Name** | **Email** | **Department** | **Institute** |
| --- | --- | --- | --- |
| Alex Bui | | Radiol Sci | UCLA |
| Antonia Petruse | | CTSI/ECRI | UCLA |
| Arash Naeim | | Medicine-Hematology/Oncology | UCLA |
| Chris Denny | | Pediatrics-Hemat & Onc | UCLA |
| Chris Denny | | Pediatrics-Hemat & Onc | UCLA |
| Clara Lajonchere | | David Geffen Sch of Med, UCLA Inst for Precision Hlth | UCLA |
| Clara Magyar | | Patholgy & Lab Med | UCLA |
| Dan Geschwind | | Neurology/Psychr & Biobehav Sci/ Human Genetics | UCLA |
| Maryam Ariannejad | | David Geffen Sch of Med, UCLA Inst for Precision Hlth | UCLA |
| Paul Boutros | | Department of Human Genetics | UCLA |
| Paul Spellman | | Medicine-Hematology/Oncology | UCLA |
| Sarah Dry | | Path & Lab Med | UCLA |
| Stan Nelson | | Hum Genetics | UCLA |
| Albert Duntugan | | UCLA Health IT \| OHIA | UCLA |
| Alex Bui | | Radiol Sci | UCLA |
| Bogdan Pasaniuc | | Genetics | Upenn |
| Chris Denny | | Ped-Hemat & Onc | UCLA |
| Clara Lajonchere | | David Geffen Sch of Med, UCLA Inst for Precision Hlth | UCLA |
| Dan Geschwind | | Neurology/Psychr & Biobehav Sci/ Human Genetics | UCLA |
| Paul Boutros | | Department of Human Genetics | UCLA |
| Paul Tung | | UCLA Health IT \| OHIA | UCLA |
| Tim Chang | | Neurology | UCLA |
| Yael Berkovich | | UCLA Health IT \| OHIA | UCLA |
| Albert Duntugan | | UCLA Health IT \| OHIA | UCLA |
| Ankur Jain | | ISS \| Ohia | UCLA |
| Danielle Martinez | | UCLA Health IT \| OHIA | UCLA |
| Ghouse Mohammed | | UCLA Health IT \| OHIA | UCLA |
| Lora Illiev | | Human Genetics | UCLA |
| Michael Broudy | | ISS OHIA | UCLA |
| Paul Boutros | | Department of Human Genetics | UCLA |
| Paul Tung | | UCLA Health IT \| OHIA | UCLA |
| Roni Haas | | Human Genetics | UCLA |
| Shaiful Alam | | Physician Support Services | UCLA |
| Taka Yamaguchi | | Jonsson Comprehensive Cancer Center/Cancer Data Center | UCLA |
| Yael Berkovich | | UCLA Health IT \| OHIA | UCLA |
| Yash Patel | | Jonsson Comprehensive Cancer Center/Cancer Data Center | UCLA |
